# Exploring Ensemble Selection for Model Averaging in Model-informed Precision Dosing: A Tacrolimus Case Study Extended to Cefepime and Infliximab

**DOI:** 10.64898/2026.09.24.26363867

**Authors:** Baroudi Samuel, Franck Bénédicte, Woillard Jean-Baptiste, Destere Alexandre, Comets Emmanuelle, Lemaitre Florian

**Affiliations:** Univ Rennes, Inserm, EHESP, Irset (Institut de recherche en santé, environnement et travail) – UMR_S 1085, F-35000 Rennes, France; FHU SUPORT, F-35000, Rennes, France; Department of Pharmacology, Toxicology and Pharmacovigilance, CHU de Limoges, Limoges, France; Pharmacology & Transplantation, INSERM U1248, Université de Limoges, Limoges, France; Department of Clinical Pharmacology, University Hospital Centre Nice Pasteur Hospital, 06000 Nice, France; Université Côte d’Azur, Inria, CNRS, Laboratoire J.A. Dieudonné, Maasai team, 06000 Nice, France; Université Paris Cité and Université Sorbonne Paris Nord, Inserm, IAME, F-75018 Paris, France

**Keywords:** population pharmacokinetics, transplantation, external evaluation, MIPD, TDM, model averaging

## Abstract

Model averaging combines the predictions of several population pharmacokinetic models to support model-informed precision dosing, but how to select ensemble models remains unexplored. Using tacrolimus as a case study, we first evaluated all possible ensembles of 27 models in 37 kidney and liver transplant recipients with full pharmacokinetic profiles at occasions 1 and 5 and trough concentrations at occasions 2, 3 and 4. Models were weighted by individual objective function value. Predictive performance for the occasion 5 area under the curve was assessed with individual parameters estimated from the occasion 5 trough concentration or from occasion 1 to 4 observations. Then, seven ensembles, selected a priori from relevance and external evaluation scores or as the best-performing ensembles previously identified, were compared across ten sets of observations, from a single trough concentration to dense sampling. Mean imprecision per ensemble size reached a plateau beyond 3-4 models. The best ensembles depended on the set of observations, comprising three models with the occasion 5 trough concentration and seven or eight models with occasions 1 to 4 observations. Across all sets, the ensemble optimised on the occasion 5 trough concentration was among the most precise, and score-based ensembles remained consistently precise, whereas the ensemble optimised on occasions 1 to 4 generalised poorly. Only a limited sampling strategy at occasion 5 kept imprecision below 20% for all ensembles. The plateau effect generalised to infliximab and cefepime. Ensemble composition matters more than ensemble size. Model averaging should rely on well-chosen ensembles rather than all available models.

## 1. Introduction

Therapeutic drug monitoring (TDM) is routinely used for drugs with a narrow therapeutic index. Model-informed precision dosing (MIPD) is increasingly integrated as a TDM tool to adapt dosing regimens by combining a population pharmacokinetic (popPK) model with individual observations. Models must also be selected carefully after assessing their relevance to the population of interest, since external evaluation studies have shown that predictive performance may not transfer reliably across populations: a model performing adequately in the cohort in which it was developed may show substantial bias when applied elsewhere. Model selection based on external evaluation studies also depends on the evaluation cohort and design, which are not consistent across studies ^1,2^. However external evaluations remain a crucial step to optimise benefit-risk ratio when using popPK models. Tacrolimus is a typical example of such a drug, given its high interindividual variability: underexposure is associated with an increased risk of acute rejection, whereas overexposure can lead to nephrotoxicity, neurotoxicity, post-transplant diabetes mellitus, and increased susceptibility to infections ^3^. Many tacrolimus popPK models have been published ^4–6^, but choosing the popPK model to use as a prior is difficult. As an alternative, popPK model averaging has emerged. In the model averaging algorithm (MAA), the predictions of several selected models are combined into a weighted average for each patient. The optimal weighting scheme has been investigated previously: weighting model predictions by each model’s individual objective function value outperformed the single model approach ^7^. However, to our knowledge, how to select models to compose the ensemble has not been investigated, and implementing all models would be impractical when numerous models have been published. In addition, the following questions therefore remain open when performing model averaging: how many models should be included to optimise ensemble performance? Does the ensemble performance increase with the number of models? Are some models redundant and interchangeable, while others are indispensable?

To answer these questions, we investigated a case study for tacrolimus model averaging. Our objective was to determine the best ensemble to implement for tacrolimus MIPD, evaluating the performance of all possible ensembles of published tacrolimus models. We also wanted to assess how accuracy and precision were impacted by the ensemble size and across different sets of observations, reflecting the sparse data available in practice across centres. Ultimately, we aimed to assess generalisability in two additional case studies for infliximab and cefepime.

## 2. Materials and Methods

### 2.1. Tacrolimus patients and data

A retrospective (2019-2025) evaluation dataset was extracted from medical records at the University Hospital of Rennes. This project was approved by the Ethics Committee of the Rennes University Hospital under approval number 25.17. The data was collected as part of routine clinical care to optimise tacrolimus dosing in a TDM context. Each patient had 5 occasions contributing 2 full-PK profiles (from 5 to 10 observations per profile) at occasion 1 and occasion 5, and 3 trough concentrations (Cmin) at occasion 2,3 and 4. In kidney transplant recipients, immunosuppressive therapy was initiated with oral tacrolimus at an initial dose of 0.1 mg/kg administered twice daily (BID). The dosage was subsequently adjusted based on TDM, targeting trough concentrations of 8–12 ng/mL from day 1 to month 3, 5–8 ng/mL thereafter, and 5–7 ng/mL beyond month 12. Patients also received mycophenolate mofetil 1 g BID and corticoids 20 mg daily, the latter dose being progressively decreased. In liver transplant recipients, the initial tacrolimus dose was 0.05 mg/kg BID (or 0.04 mg/kg in the presence of fluconazole). Mycophenolate mofetil was administered at 1.5 g BID. The tacrolimus dosing regimen was adjusted according to whole blood trough concentrations, targeting 6–10 ng/mL during the first month and 5–8 ng/mL afterwards. Patients underwent full PK sampling to estimate AUC as part of routine clinical care, with sampling schedule determined at the clinician’s discretion. Only patients with at least two available AUC estimates were included in the analysis.

Tacrolimus concentrations were determined from capillary blood samples collected using volumetric absorptive microsampling (VAMS) devices (Mitra®, Neoteryx) according to a validated high-performance liquid chromatography-tandem mass spectrometry method ^8^. The lower limit of quantification was 1 ng/mL, and intra– and inter-day precisions (CV) remained within ± 15% across the calibration range (1.0–42.9 ng/mL).

Collected covariates were sex, age, height, weight (WT), tacrolimus dose and formulation, presence and dose of corticoids, C-reactive protein (CRP), serum creatinine (SCR), diabetes status, haematocrit (HTC), haemoglobin (HB), aspartate aminotransferase (AST), post-operative days (POD) and serum albumin (ALB). Missing covariate values were imputed at the patient-occasion level using Predictive Mean Matching (PMM) via the *mice* package in R (v3.19.0) ^9^. All available covariates served as predictors, with each imputed value drawn from the d = 5 nearest-neighbour donors, and m = 10 imputed datasets were generated. Body surface area (BSA), lean body weight (LBW) and estimated glomerular filtration rate (eGFR) covariates were calculated according to equations reported in **Supporting Information**.

A dataset verification step was performed by assessing the stability of the Cmin/daily dose ratio across the 5 occasions and the distribution of POD per occasion. Patients showing a variation of at least 50% between occasion 5 and any of occasions 1 to 4 were re-investigated through re-reading of the patient’s hospital file.

### 2.2. Exhaustive evaluation of all tacrolimus model ensembles

We identified a pool of 28 tacrolimus popPK models in a previous systematic literature review ^4^. All model references are provided in **Supporting Information.** These models were implemented in Monolix®, Simulx®, and PKanalix® 2024R1 (Simulations Plus, Inc.). ^10–12^ For each model, predictions of the full PK-profile at occasion 5 were computed from the individual parameters, derived from Empirical Bayes Estimates (EBEs) by *maximum a posteriori* (MAP) estimation using i) the Cmin at occasion 5 alone, and ii) the concentrations from previous occasions 1, 2, 3, and 4. The linear-up/log-down trapezoidal method was used to compute observed area under the curve of the concentration over time for each individual *i* (*AUC_obs,i_*), and to compute for each model *m,* their corresponding predicted AUC(*AUC_pred,i,m_*).

All possible model averaging ensembles from these 28 models were assessed to evaluate the impact of ensemble size on the predictive performance of model averaging, including the single models, and ensembles from 2 to 28 models. Within each model averaging ensemble of size M, the weight of model *m* for individual *i* was calculated based on its objective function value (*OFV_m,i_*), calculated as follows ^7^:

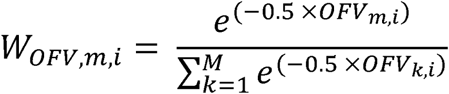

where *OFV_m,i_*, is the objective function value of model *m* for individual *i*.

The predicted AUC for individual *i*(*AUC_pred averaged,i_*) was computed as the weighted average:

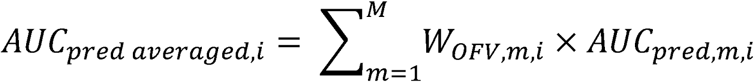

We compared and ranked model ensemble based on precision, using the relative root mean square error (RRMSE). We also computed mean prediction error (MPE) for accuracy. Both were calculated from the individual relative prediction error (*RPE_i_*), for each ensemble, as follows:

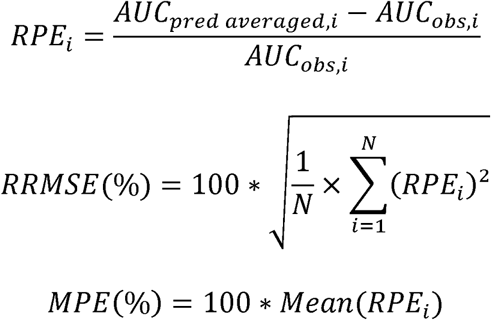

### 2.3. Effect of the set of observations for EBEs estimation on selected model ensembles

Seven model ensembles, differing in how their composition was determined, were selected for further investigation: (i) the 5 and 10 models with the highest relevance score identified in the previous systematic review ^4^ (E_RS5 and E_RS10); (ii) the 5 and 10 models with the highest external evaluation score in that review (E_EE5 and E_EE10); (iii) the 10 models with the highest combined rank across both scores (E_CR10); and (iv) the ensemble yielding the lowest RRMSE among all possible model averaging ensembles from the 28 models when EBEs were estimated using occasion 5 Cmin alone (E_Cmin) or using concentrations from occasion 1,2,3,4 (E_Multi).

Because E_Cmin and E_Multi were each optimised for one specific input scenario, whereas the five score-based ensembles were selected independently of any input scenario, we assessed how sensitive the predictive performance of all seven ensembles was to different sets of observations. Thus *AUC_pred,m,i_* at occasion 5 was computed from individual parameters obtained as EBEs by MAP estimation, using the following sets of observations: (a) a single Cmin at occasion 1, 2, 3, 4, or 5; (b) multiple Cmin at occasions 3 and 4, or at occasions 2, 3, and 4; (c) a full PK profile at occasion 1 alone, or combined with Cmin at occasions 2, 3, and 4; or (d) three observations per patient at occasion 5 selected by a limited sampling strategy (LSS) ^13^, at 0, 1, and 3 h post-dose for Prograf® and Advagraf®, and at 0, 8, and 10 h for Envarsus®. For the latter, the recommended LSS was 0, 8, and 12 hours, but the 12-hour point was replaced by a 10-hour point due to the unavailability of a 12-hour point at our centre. (**Figure 1**) ^14^.

**Figure 1.**
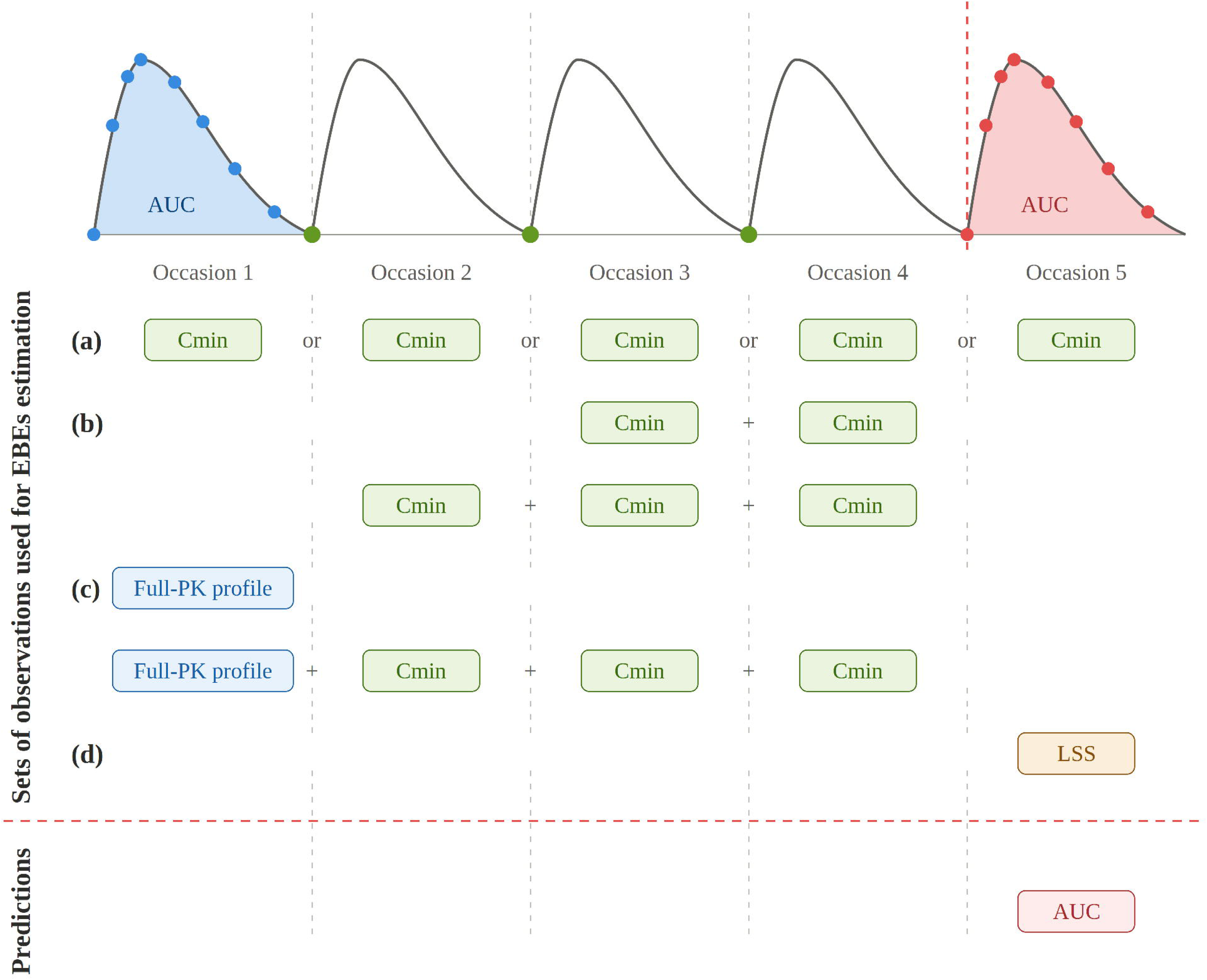
Sets of observations used for EBEs estimation to predict the AUC at occasion 5: (a) a single Cmin at one of occasions 1 to 5, (b) Cmin at occasions 3 and 4, or 2, 3 and 4, (c) the full PK profile at occasion 1, alone or combined with Cmin at occasions 2, 3 and 4, or (d) an LSS at occasion 5.

Model ensemble performances were assessed using the RRMSE and MPE for precision and accuracy, respectively (formula above). AUC were also classified as below, within, or above the therapeutic range using the transplant-specific thresholds from Ntobe-Bunkete *et al.* 2024 ^15^ (**Table S1**). Classification concordance was computed as the proportion of patients for whom *AUC_pred averaged,i_* and *AUC_obs,i_* fell within the same therapeutic category. Ultimately, we computed for each ensemble the proportion of individuals for whom the *RPE_i_* fell between –20% and +20%.

### 2.4. Complementary analyses

#### 2.4.1. Model interchangeability

First, we explored the model interchangeability within E_CR10 by quantifying the Pearson correlation coefficient of *RPE_i_* at occasion 5 using two sets of observations for EBEs estimation: 1) occasion 1,2,3,4 concentrations and 2) the Cmin at occasion 5. Two models were considered strongly correlated when the Pearson coefficient between their *RPE_i_* was ≥ 0.9. For these models, we tested whether one model could substitute another by replacing it within the model ensemble and comparing the resulting RRMSE.

#### 2.4.2. Contribution of the models from the best-performing ensemble to ensemble performance

Secondly, we compared the RRMSE of the predicted AUC at occasion 5, obtained from MAP estimation based on the Cmin at occasion 5, between all possible N-model ensembles drawn from the full set of 28 models and all possible N-model ensembles drawn from the reduced set obtained after excluding the models of E_Cmin (see definition above). This comparison determined whether comparable predictive performance could still be achieved without the models from the best ensemble.

#### 2.4.3. K-fold cross-validation

Ultimately, we performed a k-fold cross-validation using the Cmin at occasion 5, testing all possible ensembles drawn from the full set of models to assess whether model performance remained stable across folds. Patients were randomly allocated to four groups. Four folds were then defined, each using three groups as the training set and the remaining group as the test set. Within each training set, all possible model ensembles among the retained models were ranked according to their RRMSE, yielding four training ranks per ensemble, and the ensemble exhibiting the lowest mean training rank was selected as the optimal ensemble. RRMSE was also computed on the corresponding test set, and ensembles were subsequently ranked according to their mean test RRMSE (test rank).

### 2.5. Generalisability to other drugs: infliximab and cefepime case studies

We assessed generalisability to two additional drugs, cefepime and infliximab. For each drug, we performed the same exhaustive evaluation of all possible model ensembles built from the available published popPK models, applying the same weighting scheme, with concentrations predicted at one occasion from EBEs estimated using the concentrations observed at the preceding occasion(s). For the two drugs, we examined the relationship between ensemble size and RRMSE, to assess whether the pattern observed for tacrolimus was also observed for cefepime and infliximab. All methodological details are provided in **Supporting Information.**

## 3. Results

### 3.1. Tacrolimus patients and data

Thirty-seven patients were included, contributing a total of 693 observations across 5 occasions (**Table 1**). Three HTC, 4 SCR, 7 HB, 12 presence/absence and dose of corticoids, 22 diabetic status, 31 AST, 32 CRP, 40 WT, and 139 ALB values were missing and were imputed. No concentration was outside the quantification ranges. **Figure S1** shows, for each occasion, the absolute relative change in the Cmin/daily dose ratio compared with occasion 5. This change reached a mean of 33.4% between occasion 1 and occasion 5, consistent with the proximity of occasion 1 to transplantation, since occasion 5 occurred markedly later (median 367 days post-transplantation at occasion 5 vs 69 days at occasion 1, Figure S2). The change remained stable, between 20% and 25%, between occasions 2, 3, and 4 and occasion 5, occasions 2, 3, and 4 themselves being spaced one month apart. These values were considered acceptable, generally remaining below the 30% threshold frequently associated with the high inter-occasion variability of tacrolimus ^13^.

**Table 1.** Patient characteristics by occasion. OCC, occasion; AUC, area under the curve; Cmin, trough concentration; BSA, body surface area; CRP, ctive protein; AST, aspartate aminotransferase.

| Variable | OCC1 | OCC2 | OCC3 | OCC4 | OCC5 | Overall |
| --- | --- | --- | --- | --- | --- | --- |
| N patients | 37 | 37 | 37 | 37 | 37 | 37 |
| N observations | 292 | 37 | 37 | 37 | 290 | 693 |
| Continuous data – Median (min–max) |  |  |  |  |  |  |
| Cmin (ng/mL) | 9.2 (2.8–14.9) | 7.6 (2.2–11.5) | 7.5 (2.3–12.0) | 7.0 (2.7–16.4) | 6.2 (2.2–10.1) | 7.4 (2.2–16.4) |
| Daily dose (mg) | 9.5 (2–36) | 5 (1–24) | 5.0 (0.5–26.0) | 5.5 (0.5–26.0) | 5 (0.5–20) | 5.5 (0.5–36.0) |
| Age (yr) | 57 (36–72) | 57 (36–72) | 57 (36–72) | 57 (36–72) | 57 (36–72) | 57 (36–72) |
| Weight (kg) | 74.2 (50.3–126.5) | 77 (48.5–126.1) | 77 (51.0–126.1) | 77 (49.4–126.1) | 77 (50.5–145.7) | 77 (48.5–145.7) |
| Height (cm) | 172 (155–196) | 172 (155–196) | 172 (155–196) | 172 (155–196) | 172 (155–196) | 172 (155–196) |
| BSA (m²) | 1.9 (1.5–2.6) | 1.9 (1.5–2.6) | 1.9 (1.5–2.6) | 1.9 (1.5–2.6) | 1.9 (1.5–2.8) | 1.9 (1.5–2.8) |
| Haematocrit (%) | 35.3 (25.2–52.3) | 37.8 (29.5–51.7) | 37.6 (26.8–47.0) | 38.8 (30.1–49.4) | 38.7 (27.7–50) | 37.6 (25.2–52.3) |
| Post-operative days | 69 (10–242) | 181 (28–513) | 212 (53–815) | 255 (74–1208) | 367 (99–1390) | 196 (10–1390) |
| Creatinine (µmol/L) | 142 (44–229) | 126 (59–293) | 135 (52–327) | 129 (49–231) | 122 (44–226) | 132 (44–327) |
| CRP (mg/L) | 2 (0–261) | 1 (0–261) | 0 (0–261) | 0 (0–261) | 2 (0–261) | 1 (0–261) |
| AST (IU/L) | 19 (9–83) | 19 (10–278) | 20 (9–168) | 21 (10–37) | 22 (13–83) | 20 (9–278) |
| Albumin (g/L) | 39.7 (22.7–47.4) | 39.7 (30.1–47.4) | 39.7 (29.8–47.4) | 39.7 (29.8–47.4) | 39.7 (29.8–53.1) | 39.7 (22.7–53.1) |
| Haemoglobin (g/dL) | 11.3 (7.9–16.5) | 12.4 (3.6–16.9) | 12.3 (8.9–15.4) | 12.6 (9.2–15.5) | 12.2 (8.7–16.1) | 12.2 (3.6–16.9) |
| Lean body weight (kg) | 55.2 (39.1–84.2) | 55.5 (39.3–84.2) | 55.3 (39.4–84.2) | 55.3 (39.4–84.2) | 56 (39.3–92.2) | 55.3 (39.1–92.2) |
| Categorical data - N (%) |  |  |  |  |  |  |
| Hepatic transplantation | 5 (14%) | 5 (14%) | 5 (14%) | 5 (14%) | 5 (14%) | 5 (14%) |
| Renal transplantation | 30 (81%) | 30 (81%) | 30 (81%) | 30 (81%) | 30 (81%) | 30 (81%) |
| Hepatic and renal transplantation | 2 (5%) | 2 (5%) | 2 (5%) | 2 (5%) | 2 (5%) | 2 (5%) |
| Prograf (12 h) | 15 (41%) | 7 (19%) | 4 (11%) | 3 (8%) | 3 (8%) | - |
| Advagraf (24 h) | 9 (24%) | 11 (30%) | 15 (41%) | 16 (43%) | 16 (43%) | - |
| Envarsus (24 h) | 13 (35%) | 19 (51%) | 18 (49%) | 18 (49%) | 18 (49%) | - |
| Female | 17 (46%) | 17 (46%) | 17 (46%) | 17 (46%) | 17 (46%) | 17 (46%) |
| Male | 20 (54%) | 20 (54%) | 20 (54%) | 20 (54%) | 20 (54%) | 20 (54%) |
| Diabetes | 15 (41%) | 15 (41%) | 15 (41%) | 15 (41%) | 16 (43%) | - |

### 3.2. Exhaustive evaluation of all tacrolimus model ensembles

One of the 28 candidate models was excluded because its structural complexity resulted in an excessive computational time ^16^.

When estimating the EBEs with Cmin at occasion 5, the mean RRMSE of AUC at occasion 5 per ensemble size slightly decreased as the number of models included in the ensemble increased (**Figure 2**). The improvement was marginal from 39% for 3-model ensembles to 36% for the 27-model ensemble, indicating a plateau effect. For a given ensemble size, precision was highly heterogeneous among ensembles, particularly at low model counts. As the number of models increased, this variability was reduced, and the standard deviation therefore decreased. Across all ensembles, the best-performing ensemble comprised only three models (Lu *et al.* 2019, Grover *et al.* 2011, Zhu *et al.* 2014 ^17–19^), exhibiting an RRMSE of 18.7%. The mean weight and frequency of each tacrolimus model within the top 1000 ensembles is reported in **Table S2**. The Lu *et al.* 2019 ^18^ model was present in nearly all of these ensembles (97.3%), followed by Zhu *et al.* 2014 ^19^ model, in 94.5%. Lu had the highest weight reaching 21.9% while 13 models had a mean weight < 2%, even if some were useful for some patients reaching individual weight of 44.7%. Six models did not appear in any of the top 1000 ensembles. They were built predominantly on single Cmin at one or several occasions, with only a minority of patients contributing richer profiles. By contrast, the three models most frequently retained relied on markedly denser sampling schemes. Structurally, four of the 28 candidate models were described by a one-compartment structure ^20–23^; three are among the six models never selected, and the fourth contributed to only 3.9% of the top 1000 ensembles. Top 10 ensembles were composed of 2,3,4 or 5 models (**Table 2**).

**Figure 2.**
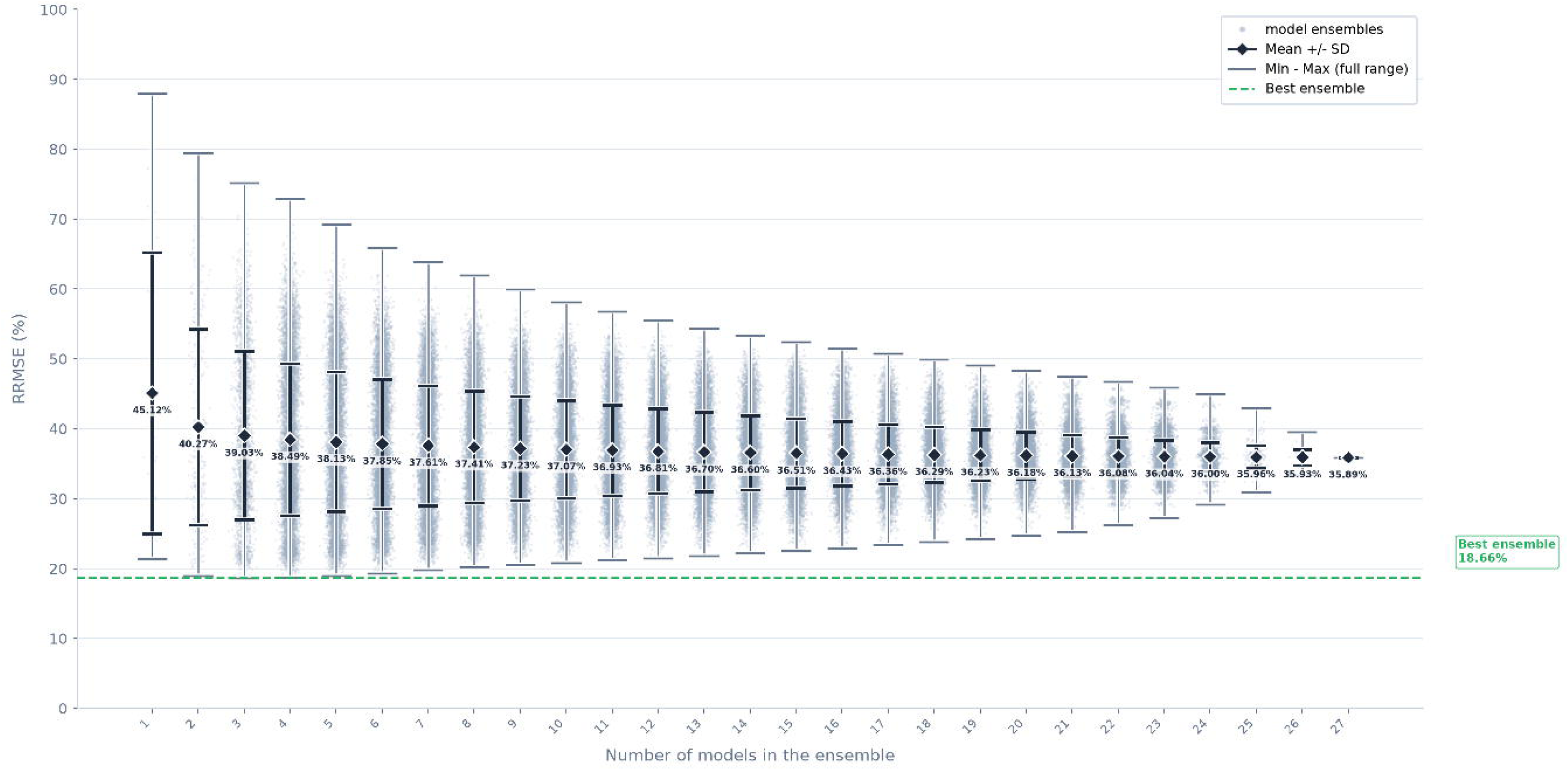
Distribution of RRMSE for the AUC at occasion 5 across all tacrolimus model ensembles grouped by ensemble size (from 1 to a maximum of 27 models per ensemble), when the occasion 5 Cmin was used for the estimation of the EBEs. The mean, standard deviation, minimum, and maximum were computed exactly over all ensembles of a given size, whereas the scatter shows a random subsample of up to 10,000 ensembles per ensemble size.

**Table 2.**
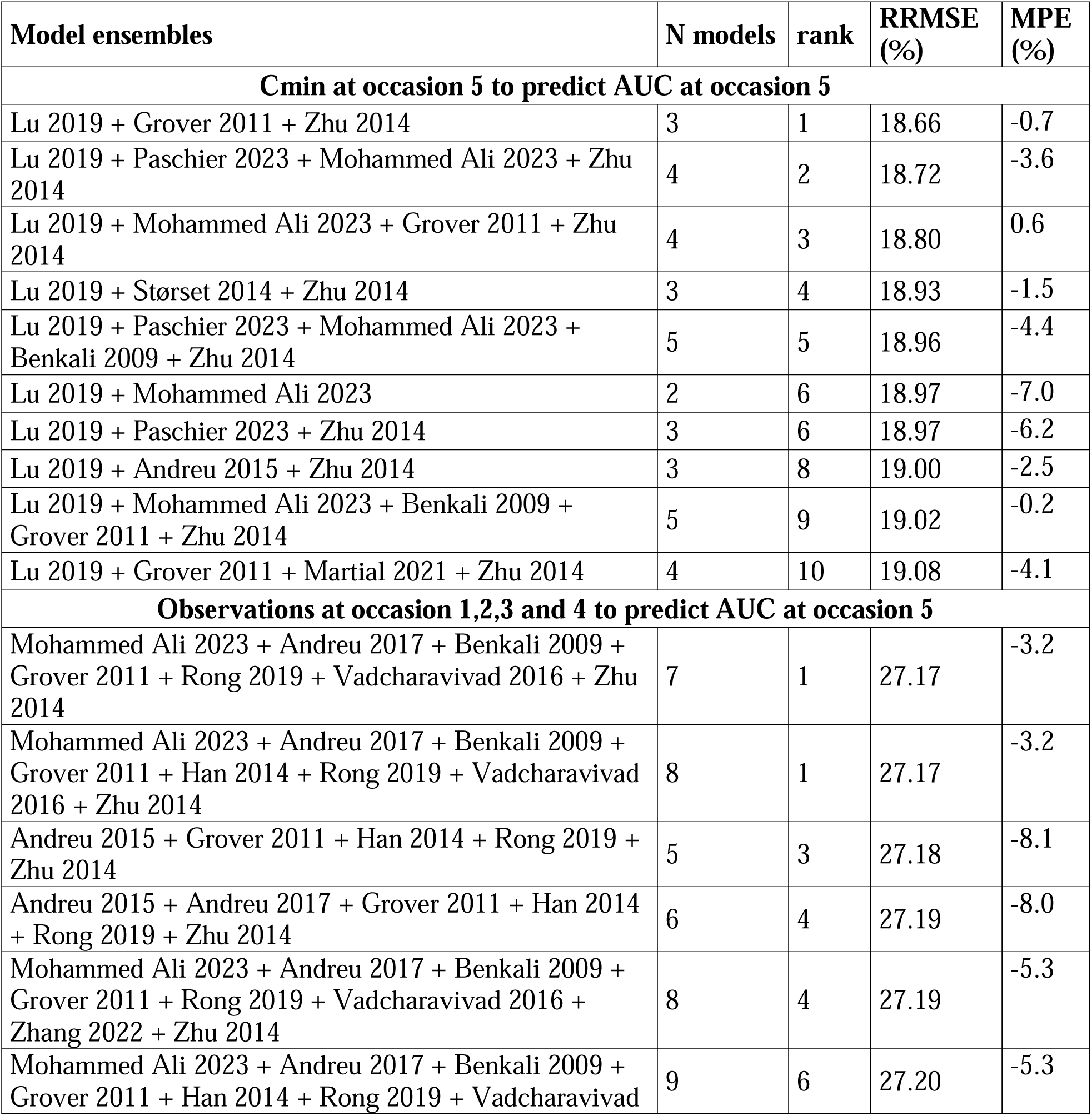
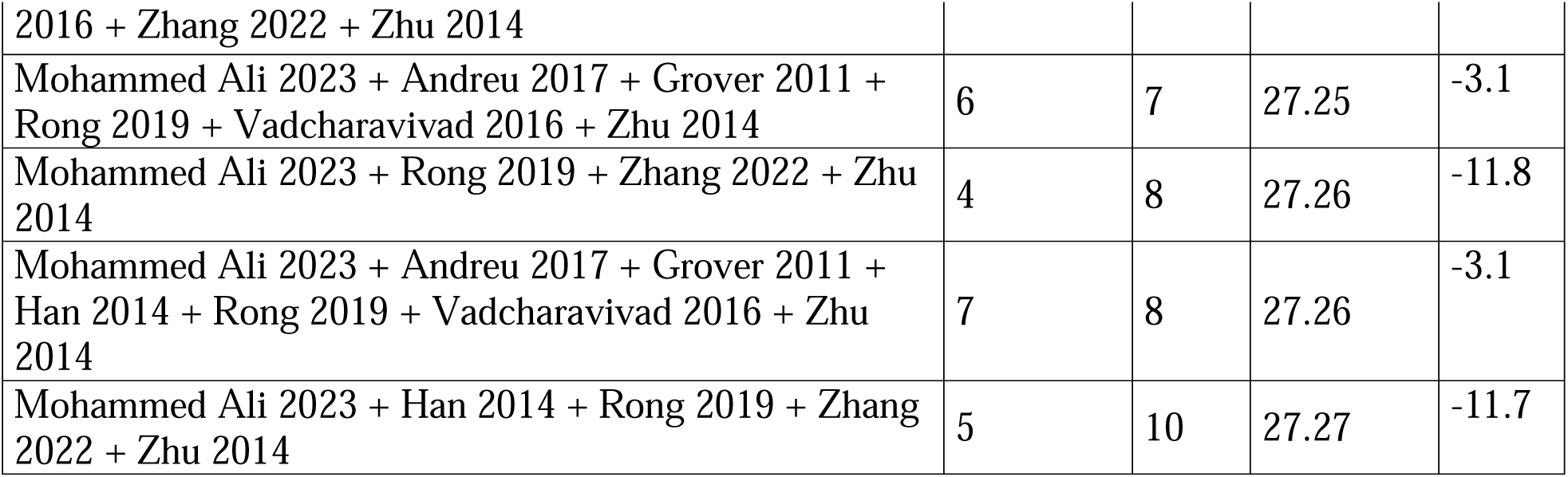
Ten ensembles with the lowest RRMSE for the AUC at occasion 5, identified through the exhaustive evaluation of tacrolimus model-averaging ensembles, when the Cmin at occasion 5 and all observations from occasions 1 to 4 were used for the estimation of the EBEs, respectively. N models: number of models within the ensemble; RRMSE, relative root mean square error; MPE, Mean prediction error. Model references are available in **Supporting Information.**

When AUC at occasion 5 was predicted using occasion 1,2,3,4 observations, the same plateau effect was found (see **Figure S3**), with a very modest improvement of the mean RRMSE with the increase of the number of models included in the ensemble. The composition of the best-performing ensembles was markedly different from that obtained with the occasion 5 Cmin: only Grover 2011 and Zhu 2014 were shared between the two best-performing ensembles, while Lu 2019, which dominated the best-performing ensemble with the occasion 5 Cmin, was absent, replaced by five additional models. The best-performing ensembles, tied at an RRMSE of 27.2%, comprised either 7 models (Mohammed Ali *et al.* 2023, Andreu *et al.* 2017, Benkali *et al.* 2009, Grover *et al.* 2011, Rong *et al.* 2019, Vadcharavivad *et al.* 2016 and Zhu *et al.* 2014 ^17,19,22,24–27^) or 8 models (the same seven plus Han *et al.* 2014 ^20^, see **Table 2**). Within the top 1000 ensembles, the frequency of each model also differed markedly from that obtained with the occasion 5 Cmin (see **Table S3**). For instance, model by Lu *et al.* 2019 was present in only 3.5% of the ensembles and its mean weight fell to 1.2%, while Zhu *et al.* 2014 was also frequently used, in 99.3% of ensembles with a mean weight of 26.9%. Performances of models also differed markedly between the two sets of observations: when used in a single model approach: Lu *et al.* 2019 or Martial *et al* 2021 ^28^ exhibited an RRMSE of 21.4% and 22.3% with occasion 5 Cmin but 48.9% and 48.1% with occasion 1,2,3,4 concentrations, respectively. (**Figure S4**)

### 3.3. Effect of the set of observations for EBEs estimation on selected model ensembles

The composition of the selected model ensembles is detailed in **Table 3**. As one model was excluded (see above), E_RS10, E_EE10 and E_CR10 comprised 9 models instead of 10. The five score-based ensembles (E_RS5, E_RS10, E_EE5, E_EE10, E_CR10) performed similarly across all sets of observations tested, regardless of the ranking criterion or ensemble size (**Figure 3**). Weight distributions across the ten sets of observations remained stable for score-based ensembles, while E_Cmin and E_Multi showed much wider weight ranges (**Figure 4**). E_Cmin achieved the best or near-best precision across nearly all sets of observations, whereas E_Multi was the least precise and the least consistent ensemble overall. Among the sets of observations, a single trough concentration collected early after transplantation (occasion 1) yielded low precision for predicting the occasion 5 AUC for every ensemble, and adding the full-PK profile at occasion 1 did not drastically improve the predictive performances. Only a limited sampling strategy at occasion 5 brought RRMSE below 20% for all ensembles, while a single Cmin collected at the occasion of the predicted AUC also gave good precision.

**Figure 3.**
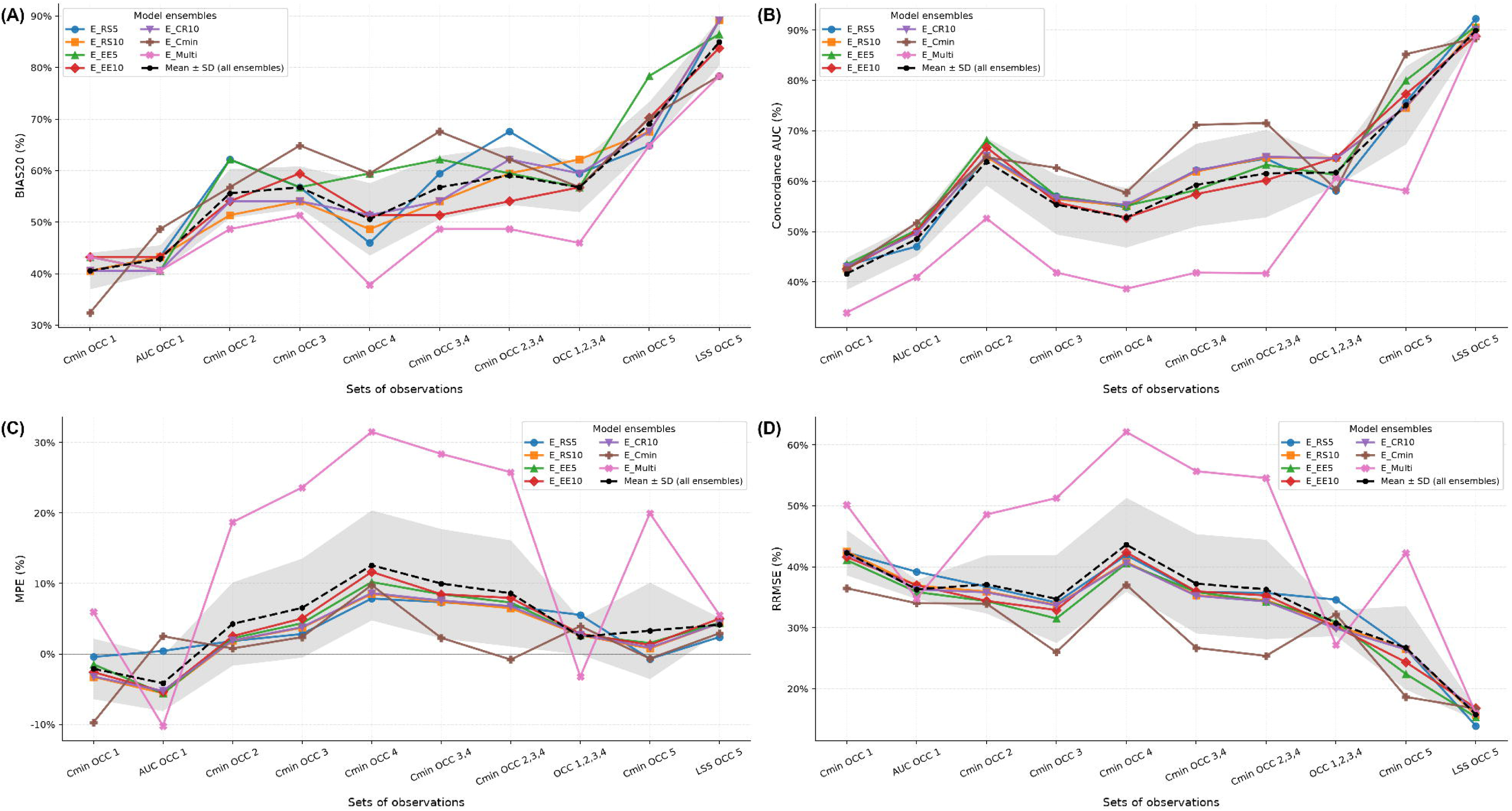
Predictive performance of the selected tacrolimus model ensembles. Each panel reports one performance metric as a function of the different sets of observations: (A) proportion of individuals for whom the fell between –20% and +20% (B) Concordance of AUC classification between predicted and observed AUC (C) Mean prediction error (MPE) (D) relative root mean square error (RRMSE). The black dashed line and shaded area represent the mean ± standard deviation (SD) of the corresponding metric across all seven ensembles, computed at each set of observations. For all curves, the line segments and shading for SD connecting adjacent sets of observations are shown for visual continuity only, as the x-axis represents discrete categories with no intermediate values.

**Figure 4.**
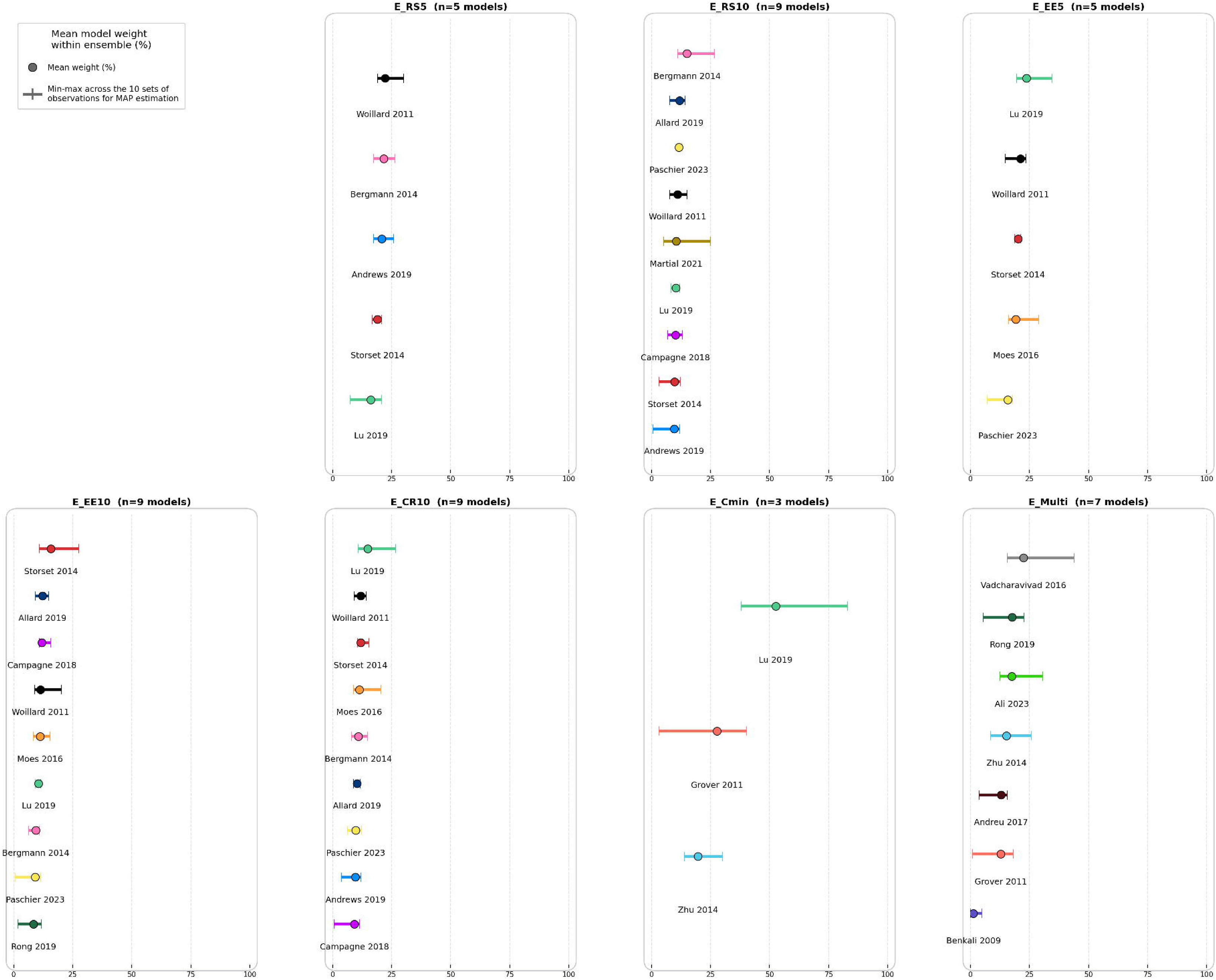
Mean weight (%) assigned to each model included within the seven selected ensembles (detailed in Table 3), with the range (min-max) across the ten sets of observations used for MAP estimation.

**Table 3.** Composition of the seven selected model ensembles. For ensembles based on the top 10 models (E_RS10, E_EE10 and E_CR10), one model was excluded because its structural complexity resulted in a computational time that would be incompatible with clinical practice resulting in 9 models ^16^. RRMSE, relative root mean square error; AUC, area under the curve. Model references are available in **Supporting Information.**

| **Model ensembles** | **Models** | **Meaning** | **N models** |
| --- | --- | --- | --- |
| E_RS5 | Andrews 2019 + Bergmann 2014 + Lu 2019 + Størset 2014 + Woillard 2011 | The 5 models with the highest relevance score. | 5 |
| E_RS10 | Allard 2019 + Andrews 2019 + Bergmann 2014 + Campagne 2018 + Lu 2019 + Paschier 2023 + Størset 2014 + Woillard 2011 + Martial 2021 | The 9 models with the highest relevance score. One model had to be excluded because it considerably increased computational time. | 9 |
| E_EE5 | Lu 2019 + Moes 2016 + Paschier 2023 + Størset 2014 + Woillard 2011 | The 5 models with the highest external evaluation scores. | 5 |
| E_EE10 | Allard 2019 + Bergmann 2014 + Campagne 2018 + Lu 2019 + Moes 2016 + Paschier 2023 + Størset 2014 + Woillard 2011 + Rong 2019 | The 9 models with the highest external evaluation scores. | 9 |
| E_CR10 | Allard 2019 + Andrews 2019 + Bergmann 2014 + Campagne 2018 + Lu 2019 + Moes 2016 + Paschier 2023 + Størset 2014 + Woillard 2011 | The 9 models with the highest combined rank across both scores. One model had to be excluded because it considerably increased computational time. | 9 |
| E_Cmin | Lu 2019 + Grover 2011 + Zhu 2014 | The ensemble yielding the lowest RRMSE among all ensembles of the 27 models for the occasion 5 Cmin to predict occasion 5 AUC. | 3 |
| E_Multi | Mohammed Ali 2023 + Andreu 2017 + Benkali 2009 + Grover 2011 + Rong 2019 + Vadcharavivad 2016 + Zhu 2014 | The ensemble yielding the lowest RRMSE among all ensembles of the 27 models for the occasion 1,2,3,4 concentrations to predict occasion 5 AUC. | 7 |

### 3.4. Complementary analyses and generalisability to other drugs

The three complementary analyses (evaluation of model interchangeability, evaluation of the contribution of the models from the best-performing ensemble to ensemble performance, and k-fold cross-validation) and generalisability assessment of the model averaging approach to infliximab and cefepime are reported in the **Supporting Information**.

In brief, for model interchangeability, substituting one correlated model (Pearson correlation coefficient of *RPE_i_* ≥0.9) for another still altered RRMSE, particularly at lower correlation coefficients (0.9-0.95), as observed when EBEs were estimated from occasion 1, 2, 3, 4 concentrations (RRMSE range 29.1-34.4%). When EBEs were instead estimated from the Cmin at occasion 5, a subset of near-perfectly correlated models (0.98-1.00) was identified that exhibited similar RRMSE values upon substitution. For the contribution of the models of the best-performing ensemble, excluding them increased mean RRMSE at every ensemble size, showing that these models contributed meaningfully to predictive performance rather than being replaceable by others. For k-fold cross-validation, the ensemble with the lowest mean training rank was also the one with the lowest mean test RRMSE (18.3%); however, the ensemble ranked first within each individual fold differed from fold to fold, even though the resulting test RRMSE values remained very close (18.4-18.8%), reflecting the presence of many near-equivalent ensembles.

Regarding generalisability to other use cases, the plateau effect observed for tacrolimus generalised to infliximab (best ensemble: 5 models, RRMSE 34.1%, plateau beyond 4 models) and cefepime (best ensemble: 2 models, RRMSE 28.7%, plateau beyond 3 models).

## 4. Discussion

In this work, we addressed the issue of selecting a set of models to include in a model averaging ensemble, focusing on tacrolimus. We assessed the optimal number of models in an ensemble, whether some models are redundant or indispensable, and the generalisability of our findings. On average, adding models brought only marginal gains beyond 3 to 4 models for tacrolimus, a plateau that generalised to the cefepime and infliximab use cases. This plateau, however, only describes the average trend by ensemble size. The best combination identified achieved an RRMSE of 18.7% with individual parameters derived from Cmin at occasion 5, against 35.9% for the full pool of 27 models; ensembles built from our relevance and external evaluation scores achieved RRMSE between 22% and 27%, a marked improvement over including all candidate models. Despite this plateau effect, models were not always interchangeable. We tested it with E_CR10, and prediction errors were correlated (Pearson coefficient ≥0.9) between some tacrolimus models, but only near-perfect correlation made models completely interchangeable, so a 0.9 threshold is too low to identify substitutable models. The best ensemble was also not stable across patients: in the k-fold analysis, the top-ranked ensembles differed between training folds, although their test RRMSE remained similar, showing that many model ensembles had near-equivalent precision and that no single best ensemble could be identified. But excluding the models from the best-performing ensemble from the pool of candidate models lowered the mean precision at every ensemble size, confirming that some models remained essential contributors. Of note, the model by Grover *et al.* was the least precise model in the single-model approach with the occasion 5 Cmin (RRMSE 88.0%), yet it belonged to the best-performing ensemble, suggesting that the contribution of a model to an ensemble depends on how its prediction errors complement those of the other models, rather than on its standalone performance.

Thus, no single ensemble size or composition emerged as universally optimal. Score-based ensembles, however, consistently ranked among the most precise across all sets of observations tested, unlike E_Multi, tailored to one specific scenario, which generalised poorly. Building an ensemble from a relevance or external evaluation score is therefore a valuable strategy (depending on the availability of a dataset for evaluation) and remains preferable to an arbitrary selection of models to average, since it approximates exhaustive search results without its computational cost.

For our tacrolimus use case, the marked differences observed between occasions 1,2,3,4 and Cmin at occasion 5 could be explained by the highly variable pharmacokinetics of the drug during the early post-transplant period (median POD 69 vs 367 days at occasions 1 and 5) ^29^. This implies further investigation in additional use cases, ideally with a direct comparison of different sets of observations for EBEs estimation, to determine whether this dependence on the set of observations is specific to tacrolimus or applies more broadly. Practically, these differences in model performance across sets of observations highlighted that, for the implementation of an MIPD strategy, the optimal performance was achieved with the Cmin at occasion 5 and the occasion 5 LSS, both offering performance suitable for clinical use, unlike sets of observations based on a single, more distant Cmin or a distant AUC. An MIPD strategy could therefore be implemented with these sets of observations to estimate individual parameters, combining high accuracy and high precision.

This study has limitations: all three use cases relied on a single-centre dataset with a limited number of patients. Thus, tacrolimus k-fold analysis should be interpreted cautiously given the cohort size (n = 37). Also, ensembles were ranked by RRMSE alone, and only the OFVi-based weighting scheme was explored. In addition, the Cmin/daily dose ratio showed substantial inter-occasion variability, particularly at occasion 1, which is close to transplantation. Patients with the largest variations were re-investigated using the information available in their hospital records, but the retrospective design of the study did not allow further verification. Additionally, a substantial proportion of albumin values were missing (75%) and were imputed using PMM. As only 2 models incorporated albumin as a covariate, and comparing results across several datasets with different albumin imputations would have required re-running the exhaustive combinatorial evaluation, which was computationally too costly, a sensitivity analysis was not performed. Finally, tacrolimus popPK models from literature were developed in heterogeneous transplant populations. This heterogeneity may constitute an additional source of variability between model predictions, an aspect that was not specifically investigated here.

To conclude, no single optimal number of models for model averaging emerged across the three use cases. Ensemble performance did not keep increasing with the number of models: a plateau was reached early, and including more models than needed brought no further benefit but did not compromise precision on average either. Models were interchangeable only when their prediction errors were near-perfectly correlated, while others carried information that could not be replaced. Overall, model averaging should not be reduced to combining as many published models as possible: the design of the ensemble matters. This work suggests that a subset of well-chosen models, selected with prior validated score, can approach the performance of the best-performing models at a fraction of the computational cost of an exhaustive evaluation. We therefore identified a well-performing ensemble and sets of observations suitable for tacrolimus MIPD. Future work should link model characteristics, such as structure and sampling design, to their contribution to ensemble precision, and confirm these findings in a larger, multicentre cohort.

## Study Highlights

### What is the current knowledge on the topic?

Model averaging, in which popPK model predictions are weighted by their individual objective function value, improves the predictive performance of MIPD compared with a single-model approach. However, for drugs such as tacrolimus, for which numerous popPK models have been published, no guidance exists on how to compose the ensemble.

### What question did this study address?

How many and which models should be included in a model averaging ensemble? Are some models redundant and interchangeable, or indispensable? Do these findings generalise beyond tacrolimus?

### What does this study add to our knowledge?

The exhaustive evaluation of all possible tacrolimus ensembles showed that precision reached a plateau beyond 3-4 models, a pattern also observed for infliximab and cefepime. A small, well-chosen ensemble outperformed the full pool of 27 models (RRMSE 18.7% vs 35.9%), but the optimal composition depended on the set of observations used for individual parameters estimation. Models were interchangeable only when their prediction errors were near-perfectly correlated, and some models could not be replaced.

### How might this change clinical pharmacology or translational science?

Optimising model averaging does not mean combining as many published models as possible. Selecting a limited ensemble based on relevance or external evaluation scores approximates exhaustive search at a fraction of its computational cost and remains robust across sets of observations available for individual parameters estimation.

## 5. Acknowledgments

Authors would like to thank information specialists Damien Belvèze and Xavier Chard-Hutchinson for their peer review of the search strategy following the PRESS methodology for infliximab and tacrolimus case studies, Florian Naudet for his contributions regarding systematic literature review, Sylvain Benito and Julien Welzel for their shared opinions regarding model averaging.

## 6. Authors Contributions

SB wrote the manuscript; BF, EC, FL, JBW, SB designed the research; AD, BF and SB performed the research; SB analyzed the data.

## Data Availability

All data produced in the present study are available upon reasonable request to the authors.

## 10. Supporting Information titles

1. Supporting information regarding tacrolimus model averaging
  1.1. Methods
    1.1.1. Tacrolimus patients and data
    1.1.2. Model references
  1.2. Additional results regarding tacrolimus
    1.2.1. Complementary analyses
      1.2.1.1. Model interchangeability
      1.2.1.2. Contribution of the models from the best-performing ensemble to ensemble performance
      1.2.1.3. K-fold cross-validation
2. Generalisability to other drugs: infliximab model averaging
  2.1. Method
    2.1.1. Infliximab patients and data
      2.1.1.1. Systematic literature review of infliximab models
    2.1.2. Evaluation design and exhaustive evaluation of all infliximab model ensembles
  2.2. Results
3. Generalisability to other drugs: cefepime model averaging
  3.1. Method
    3.1.1. Literature review, data collection, model implementation, patients and data
    3.1.2. Evaluation design and exhaustive evaluation of all cefepime model ensembles
  3.2. Results
4. Supplementary Figures and tables.
  - **Table S1**. Therapeutic range for Tacrolimus AUC.
  - **Figure S1.** Absolute relative change in Cmin/daily dose ratio relative to occasion 5 for each previous occasion. Results show absolute median and absolute mean ± standard deviation.
  - **Figure S2.** Post-operative days distribution for each occasion.
  - **Table S2.** Mean, minimum, and maximum weight (%) of each tacrolimus model within the top 1000 ensembles when the Cmin at occasion 5 was used for the estimation of the EBEs to predict the AUC at occasion 5, and frequency of each model among the top 1000 ensembles.
  - **Figure S3.** Distribution of RRMSE across all tacrolimus model ensembles grouped by ensemble size (from 1 to a maximum of 27 models per ensemble), when the occasion 1, 2, 3, 4 observations were used for the estimation of the EBEs to predict the occasion 5 AUC. The mean, standard deviation, minimum, and maximum were computed exactly over all ensembles of a given size, whereas the scatter shows a random subsample of up to 10,000 ensembles per ensemble size.
  - **Table S3.** Mean, minimum, and maximum weight (%) of each tacrolimus model within the top 1000 ensembles using occasion 1,2,3,4 observations to predict occasion 5 AUC, and frequency of each model among the top 1000 ensembles.
  - **Figure S4.** Single model approach evaluation assessing the RRMSE for each model for the two sets of observations evaluated (Cmin at occasion 5 and observations at occasions 1,2,3 and 4 to AUC at occasion 5).
  - **Figure S5.** Pairwise Pearson correlation of occasion 5 AUC individual relative prediction error across the 9 tacrolimus popPK models from E_CR10. Occasion 5 AUC predicted from EBEs estimated with (A) the concentrations from occasions 1, 2, 3, 4, and (B) the Cmin at occasion 5.
  - **Table S4.** RRMSE (%) of model ensembles combining a fixed base set with one interchangeable model drawn from a correlated group, for the two sets of observations used for the estimation of the EBEs (occasion 1, 2, 3, 4 concentrations and occasion 5 Cmin).
  - **Figure S6.** Distribution of RRMSE (%) across all tacrolimus model ensembles as a function of ensemble size, with and without models from the best-performing ensemble (Lu 2019, Grover 2011, and Zhu 2014), using Cmin at occasion 5.
  - **Table S5.** Top ten of ensembles with the lowest mean training rank in the K-fold cross-validation. N models: number of models within the ensemble; RRMSE, relative root mean square error; SD, standard deviation.
  - **Table S6.** Optimal model ensemble selected on the mean training rank and ensembles ranked first within each individual fold. RRMSE, relative root mean square error.
  - **Table S7.** Patient and dosing characteristics. Infliximab PK database (stratified by occasion).
  - **Figure S7.** Prisma flow chart for infliximab systematic literature review.
  - **Table S8.** Patient characteristics and covariate distribution for cefepime dataset.
  - **Figure S8.** Distribution of RRMSE across all model ensembles grouped by ensemble size (from 1 to a maximum of 14 models per ensemble), for infliximab use case.
  - **Table S9.** Top ten ensembles with the lowest RRMSE, identified through exhaustive evaluation of infliximab model-averaging ensembles. N models: number of models within the ensemble; RRMSE, relative root mean square error.
  - **Table S10.** Mean, minimum, and maximum weight (%) of each infliximab model within the top 1000 ensembles for OCC5 Cmin prediction based on OCC1,2,3,4 Cmin, and frequency of each model among the top 1000 ensembles.
  - **Figure S9.** Distribution of RRMSE across all model ensembles grouped by ensemble size (from 1 to a maximum of 9 models per ensemble), for cefepime use case.
  - **Table S11.** Top ten ensembles with the lowest RRMSE, identified through exhaustive evaluation of cefepime model-averaging ensembles. N models: number of models within the ensemble; RRMSE, relative root mean square error.
  - **Table S12**. Mean, minimum, and maximum weight (%) of each cefepime model within top 30 ensembles for OCC2 concentrations prediction based on OCC1 concentrations, and frequency of each model among top 30 ensembles.

## References

1. Cheng Y, Wang C yu, Li Z ran, Pan Y, Liu M bai, Jiao Z. Can Population Pharmacokinetics of Antibiotics be Extrapolated? Implications of External Evaluations. Clin Pharmacokinet. 2021;60(1):53–68. doi:10.1007/s40262-020-00937-4

2. El Hassani M, Marsot A. External Evaluation of Population Pharmacokinetic Models for Precision Dosing: Current State and Knowledge Gaps. Clin Pharmacokinet. 2023;62(4):533–540. doi:10.1007/s40262-023-01233-7

3. Destere A. Suivi du patient transplanté rénal traité par tacrolimus par pharmacocinétique de population et machine learning. phdthesis. Université de Limoges; 2022. Accessed June 19, 2025. https://theses.hal.science/tel-04402665

4. Baroudi S, Franck B, Woillard JB, Béni NB, Comets E, Lemaitre F. Relevance-Based Score for MIPD Model Selection: Systematic Review and External Evaluation of popPK Tacrolimus Models in Adult Transplant Recipients. CPT Pharmacometrics Syst Pharmacol. 2026;15(8):e70315. doi:10.1002/psp4.70315

5. Kirubakaran R, Stocker SL, Hennig S, Day RO, Carland JE. Population Pharmacokinetic Models of Tacrolimus in Adult Transplant Recipients: A Systematic Review. Clin Pharmacokinet. 2020;59(11):1357–1392. doi:10.1007/s40262-020-00922-x

6. Zhao CY, Jiao Z, Mao JJ, Qiu XY. External evaluation of published population pharmacokinetic models of tacrolimus in adult renal transplant recipients. Br J Clin Pharmacol. 2016;81(5):891–907. doi:10.1111/bcp.12830

7. Uster DW, Stocker SL, Carland JE, et al. A Model Averaging/Selection Approach Improves the Predictive Performance of Model-Informed Precision Dosing: Vancomycin as a Case Study. Clin Pharmacol Ther. 2021;109(1):175–183. doi:10.1002/cpt.2065

8. Tron C, Ferrand-Sorre MJ, Querzerho-Raguideau J, et al. Volumetric absorptive microsampling for the quantification of tacrolimus in capillary blood by high performance liquid chromatography-tandem mass spectrometry. J Chromatogr B Analyt Technol Biomed Life Sci. 2021;1165:122521. doi:10.1016/j.jchromb.2020.122521

9. Buuren S van, Groothuis-Oudshoorn K. mice: Multivariate Imputation by Chained Equations in R. Journal of Statistical Software. 2011;45:1–67. doi:10.18637/jss.v045.i03

10. Monolix 2024 R1, Simulations Plus, doi: 10.5281/zenodo.11401936.

11. Simulx 2024 R1, Simulations Plus, doi: 10.5281/zenodo.11402004.

12. PKanalix 2024 R1, Simulations Plus, doi: 10.5281/zenodo.11401684.

13. Woillard JB, de Winter BCM, Kamar N, Marquet P, Rostaing L, Rousseau A. Population pharmacokinetic model and Bayesian estimator for two tacrolimus formulations--twice daily Prograf and once daily Advagraf. Br J Clin Pharmacol. 2011;71(3):391–402. doi:10.1111/j.1365-2125.2010.03837.x

14. Woillard JB, Debord J, Monchaud C, Saint-Marcoux F, Marquet P. Population Pharmacokinetics and Bayesian Estimators for Refined Dose Adjustment of a New Tacrolimus Formulation in Kidney and Liver Transplant Patients. Clin Pharmacokinet. 2017;56(12):1491–1498. doi:10.1007/s40262-017-0533-5

15. Ntobe-Bunkete B, Lemaitre F. Therapeutic drug monitoring in kidney and liver transplantation: current advances and future directions. Expert Rev Clin Pharmacol. 2024;17(5-6):505–514. doi:10.1080/17512433.2024.2354276

16. Henin E, Govoni M, Cella M, Laveille C, Piotti G. Therapeutic Drug Monitoring Strategies for Envarsus in De Novo Kidney Transplant Patients Using Population Modelling and Simulations. Adv Ther. 2021;38(10):5317–5332. doi:10.1007/s12325-021-01905-5

17. Grover A, Frassetto LA, Benet LZ, Chakkera HA. Pharmacokinetic differences corroborate observed low tacrolimus dosage in Native American renal transplant patients. Drug Metab Dispos. 2011;39(11):2017–2019. doi:10.1124/dmd.111.041350

18. Lu Z, Bonate P, Keirns J. Population pharmacokinetics of immediate– and prolonged-release tacrolimus formulations in liver, kidney and heart transplant recipients. Br J Clin Pharmacol. 2019;85(8):1692–1703. doi:10.1111/bcp.13952

19. Zhu L, Wang H, Sun X, et al. The Population Pharmacokinetic Models of Tacrolimus in Chinese Adult Liver Transplantation Patients. J Pharm (Cairo). 2014;2014:713650. doi:10.1155/2014/713650

20. Han N, Ha S, Yun H yeol, et al. Population pharmacokinetic-pharmacogenetic model of tacrolimus in the early period after kidney transplantation. Basic Clin Pharmacol Toxicol. 2014;114(5):400–406. doi:10.1111/bcpt.12176

21. Staatz CE, Willis C, Taylor PJ, Lynch SV, Tett SE. Toward better outcomes with tacrolimus therapy: population pharmacokinetics and individualized dosage prediction in adult liver transplantation. Liver Transpl. 2003;9(2):130–137. doi:10.1053/jlts.2003.50023

22. Vadcharavivad S, Praisuwan S, Techawathanawanna N, Treyaprasert W, Avihingsanon Y. Population pharmacokinetics of tacrolimus in Thai kidney transplant patients: comparison with similar data from other populations. J Clin Pharm Ther. 2016;41(3):310–328. doi:10.1111/jcpt.12396

23. Teng F, Zhang W, Wang W, et al. Population pharmacokinetics of tacrolimus in Chinese adult liver transplant patients. Biopharm Drug Dispos. 2022;43(2):76–85. doi:10.1002/bdd.2311

24. Mohammed Ali Z, Meertens M, Fernández B, et al. CYP3A5*3 and CYP3A4*22 Cluster Polymorphism Effects on LCP-Tac Tacrolimus Exposure: Population Pharmacokinetic Approach. Pharmaceutics. 2023;15(12):2699. doi:10.3390/pharmaceutics15122699

25. Andreu F, Colom H, Elens L, et al. A New CYP3A5*3 and CYP3A4*22 Cluster Influencing Tacrolimus Target Concentrations: A Population Approach. Clin Pharmacokinet. 2017;56(8):963–975. doi:10.1007/s40262-016-0491-3

26. Benkali K, Prémaud A, Picard N, et al. Tacrolimus population pharmacokinetic-pharmacogenetic analysis and Bayesian estimation in renal transplant recipients. Clin Pharmacokinet. 2009;48(12):805–816. doi:10.2165/11318080-000000000-00000

27. Rong Y, Mayo P, Ensom MHH, Kiang TKL. Population Pharmacokinetic Analysis of Immediate-Release Oral Tacrolimus Co-administered with Mycophenolate Mofetil in Corticosteroid-Free Adult Kidney Transplant Recipients. Eur J Drug Metab Pharmacokinet. 2019;44(3):409–422. doi:10.1007/s13318-018-0525-3

28. Martial LC, Biewenga M, Ruijter BN, et al. Population pharmacokinetics and genetics of oral meltdose tacrolimus (Envarsus) in stable adult liver transplant recipients. Br J Clin Pharmacol. 2021;87(11):4262–4272. doi:10.1111/bcp.14842

29. Sikma MA, Hunault CC, Van Maarseveen EM, et al. High Variability of Whole-Blood Tacrolimus Pharmacokinetics Early After Thoracic Organ Transplantation. Eur J Drug Metab Pharmacokinet. 2020;45(1):123–134. doi:10.1007/s13318-019-00591-7

