## Supporting Information for "Exploring Ensemble Selection for Model Averaging in Model-informed Precision Dosing: A Tacrolimus Case Study Extended to Cefepime and Infliximab"

### 1. Supporting information regarding tacrolimus model averaging

#### 1.1. Methods

##### 1.1.1. Tacrolimus patients and data

Additional required covariates were calculated as follows:

- Body surface area (BSA) in m² from Mosteller *et al.* 1987 formula ^1^:

BSA = $\sqrt{\frac{height\left( cm \right)\times weight(kg)}{3600}}$

- Lean body weight (LBW) in kg from Boer *et al* ^2^*.*

$$LBW_{males}=0.407\times weight \left( kg \right)+0.267\times height (cm)-19.2$$

$$LBW_{females}=0.252\times weight \left( kg \right)+0.473\times height (cm)-48.3$$

- Estimated glomerular filtration rate (eGFR) in mL/min/1.73 m² with MDRD formula from Levey *et al.* ^3^:

$${eGFR}_{MDRD}=175\times{(\frac{creat}{88.4})}^{-1.154}\times{age}^{-0.203}\times(0.742 if female)$$

- eGFR in mL/min with Cockcroft formula from Cockcroft and Gault ^4^:

$${eGFR}_{{cockcroft}_{males}}=1.23\times weight\times\frac{(140-age)}{creat}$$

$${eGFR}_{{cockcroft}_{females}}=1.04\times weight\times\frac{(140-age)}{creat}$$

Where creat is in μmol/L, $age$ is in years, $weight$ in kg.

##### 1.1.2. Model references

The pool of 28 tacrolimus popPK models included Allard *et al.*, 2019, Andreu *et al.*, 2015, Andreu *et al.*, 2017, Andrews *et al.*, 2019, Benkali *et al.*, 2009, Benkali *et al.*, 2010, Bergmann *et al.*, 2014, Campagne *et al.*, 2018, Grover *et al.*, 2011, Han *et al.*, 2014, Henin *et al.*, 2021, Kamp *et al.*, 2023, Lu *et al.*, 2019, Martial *et al.*, 2021, Moes *et al.*, 2016, Mohammed Ali *et al.*, 2023, Ogasawara *et al.*, 2013, Paschier *et al.*, 2023, Press *et al.*, 2009, Rong *et al.*, 2019, Sikma *et al.*, 2020, Staatz *et al.*, 2003, Størset *et al.*, 2014, Teng *et al.*, 2022, Vadcharavivad *et al.*, 2016, Woillard *et al.*, 2011, Zhang *et al.*, 2022, and Zhu *et al.*, 2014. ^5–32^

#### 1.2. Additional results regarding tacrolimus

##### 1.2.1. Complementary analyses

###### 1.2.1.1. Model interchangeability

The Pearson correlation coefficients of ${RPE}_{i}$ at occasion 5 across all models are shown in **Figure S5**. When the occasion 1, 2, 3, 4 observations were used for the estimation of the EBEs, three groups emerged: two models (Campagne 2018 and Lu 2019) showed correlation coefficients below 0.9 with all other models, and one model (Bergmann 2014) showed coefficients close to 0.9, though some fell below this threshold, whereas the six remaining models were mutually correlated with coefficients around 0.9. RRMSE of ensembles combining these four models with each model drawn from the correlated group (**Table S4**) ranged from 29.1 to 34.4%, exhibiting different predictive performance even when strongly correlated. When the Cmin at occasion 5 was used instead, a group of four models (Allard 2019, Moes 2016, Campagne 2018, and Paschier 2023) emerged with even stronger mutual correlations than with the first set of observations, with coefficients ranging from 0.98 to 1. The RRMSE of ensembles combining a fixed set of five models (Lu 2019, Størset 2014, Bergmann 2014, Andrews 2019, and Woillard 2011) with one model drawn from this highly correlated group was then compared (**Table S4**). The RRMSE range (26.4 to 27.7%) was narrower than under the first set of observations, consistent with the higher Pearson correlation coefficients observed among the tested models.

###### 1.2.1.2. Contribution of the models from the best-performing ensemble to ensemble performance

For every ensemble size, the ensembles drawn from the reduced set exhibited higher RRMSE relative to that of ensembles drawn from the 27 models, indicating a consistent loss of predictive performance once models from the best-performing ensemble were excluded from selection (**Figure S6)**. The lowest RRMSE attainable from the reduced set was 21.9%, compared with 18.7% for the full set, and this gap persisted across all ensemble sizes. The mean RRMSE of the reduced-set ensembles remained higher at every ensemble size.

###### 1.2.1.3. K-fold cross-validation

The 37 patients were randomly allocated into four groups (A: n = 10; B: n = 9; C: n = 9; D: n = 9). The ensemble achieving the lowest mean training rank (16.25) was a four-model ensemble comprising Lu 2019, Paschier 2023, Mohammed Ali 2023 and Zhu 2014 ^17,20,22,32^. This ensemble yielded test RRMSE values of 19.87%, 20.06%, 12.26% and 21.21% in folds A to D, corresponding to a mean test RRMSE of 18.3% (SD 4.1%), which was also the lowest value observed among all ensembles (**Table S5**). Selection based on the mean training rank therefore identified the same ensemble as direct ranking on the test sets. The ten ensembles with the lowest mean training ranks contained 2 to 5 models (mean test RRMSE ranging from 18.3% to 18.9%). Lu 2019 was present in all ten and Zhu 2014 in nine, consistent with the exhaustive evaluation, in which these models were present in almost all of the top 1000 ensembles. Conversely, the ensembles ranked first within a single fold showed markedly less stability across folds, with mean training ranks ranging from 47.5 to 4,732.8 (**Table S6**). However, their mean test RRMSE values ranged from 18.4% to 18.8%, reflecting the density of near-equivalent ensembles rather than a loss of predictive performance across folds.

### 2. Generalisability to other drugs: infliximab model averaging

#### 2.1. Method

##### 2.1.1. Infliximab patients and data

A retrospective (2012-2021) evaluation dataset was extracted from medical records at the University Hospital of Rennes. 187 patients contributed Cmin on 5 occasions constituting a total of 935 occasions (**Table S7**). Collected covariates were sex, age, weight, dose, infusion duration, C-reactive protein (CRP), calprotectin, HBI (Harvey Bradshaw Index), PDAI (Perianal Disease Activity Index), serum albumin (ALB), and co-treatments (Methotrexate, Thioguanine, Azathioprine, Mercaptopurine). A binary maintenance covariate (yes/no) was defined based on time since treatment initiation: patients were classified as "maintenance" if infliximab was initiated more than 6 weeks prior, and as "initiation" otherwise, following Buurman *et al.* 2015 ^33^. No covariate value was missing.

The concentrations were collected as part of routine clinical care to optimise infliximab dosing in a TDM context. Bioassay was performed according to a previously published method. ^34^ Dosage at initiation and subsequent dose adaptation was already described elsewhere. ^35^

The local ethic committee approved the protocol. Patients included in the study gave their consent to be registered in a database for research use, and the study was approved by the “Commission Nationale Informatique et Liberté” (CNIL N°1412467).

##### 2.1.2. Systematic literature review of infliximab models

A systematic literature review was performed to identify infliximab popPK models in patients with inflammatory bowel disease (IBD). The review was conducted following PRISMA 2020 guidelines ^36^. The complete protocol was registered on PROSPERO (registration number CRD42024519679) prior to the start of data collection. Medline was accessed through the PubMed® platform, and Web of Science was used as a complementary database. The PubMed® search query was peer-reviewed by an information specialist following the PRESS methodology ^37^, resulting in the addition of a MeSH term to restrict the search to the IBD. The following query was used:

("Inflammatory Bowel Diseases"[Mesh]) AND ("infliximab" AND ("population pharmacokinetic*" OR "pharmacokinetic* model*" OR "popPK model*" OR "popPK" OR "PK model*")). The same query was adapted and applied to Web of Science. The search was initiated in March 2024, with ongoing state-of-the-art monitoring performed through an RSS feed managed in Zotero until August 27, 2026. Publications in languages other than English or French were excluded. Records identified from PubMed (n=74) and Web of Science (n=126) were pooled (n=200); 56 duplicate records were removed before screening (n=144). Two authors (SB and AD) independently screened titles and abstracts against the eligibility criteria, excluding records concerning other drugs, other populations, non-popPK models, or unpublished studies (n=117 excluded), yielding 27 reports sought for retrieval (see **Figure S7**). Nine studies conducted in children or pregnant women were excluded per the exclusion criteria, leaving 18 reports assessed for eligibility. Four reports were further excluded (2 not reproducible, 2 with no access to the complete original publication), resulting in 14 studies included in the review.

Included infliximab popPK models were: Berends *et al.* 2019, Brandse *et al.* 2016, Brandse *et al.* 2017, Buurman *et al.* 2015, Dotan *et al.* 2014, Dreesen *et al.* 2021, Edlund *et al.* 2017, Fasanmade *et al.* 2011, Fasanmade *et al.* 2009, Grisic *et al.* 2021, Kantasiripitak *et al.* 2021, Magro *et al.* 2024, Passot *et al.* 2016, and Ternant *et al.* 2018. ^33,38–50^

##### 2.1.3. Evaluation design and exhaustive evaluation of all infliximab model ensembles

Models were implemented on Monolix® and Simulx®. 2024R1 (Simulations Plus, Inc.). ^51,52^ For the 14 models, Cmin at occasion 5 were computed from the individual parameters derived from EBEs by MAP estimation using Cmin observations from occasion 1 to 4. For model averaging, we applied the same weighting scheme as for tacrolimus, based on the OFV ($W_{OFV,m,i}$). For each model ensemble, RRMSE was computed as an imprecision metric to evaluate all infliximab model averaging ensembles as follows:

$${RPE}_{i}=\frac{{Cmin}_{pred averaged,i}-{Cmin}_{obs,i}}{{Cmin}_{obs,i}}$$

$$RRMSE(\text{\%})=100*\sqrt{\frac{1}{N}\times\sum_{i=1}^{N} \left( {RPE}_{i} \right)^{2}}$$

#### 2.2. Results

The mean RRMSE per ensemble size decreased as more models were included in model ensembles, with only marginal further improvement beyond four models (from 36.8% for 4-model ensembles to 36% for the full 14-model ensemble), consistent with a plateau effect (**Figure S8**). Variability was greater for small ensembles and narrowed progressively as more models were included. Across all ensembles, the best-performing ensemble comprised five models (Edlund *et al.* 2017, Fasanmade *et al.* 2011, Kantasiripitak *et al.* 2021, Magro *et al.* 2024 and Passot *et al.* 2016), with an RRMSE of 34.1% (**Table S9**). The ten best-performing ensembles included from 3 to 7 models (RRMSE 34.1-34.3%), all including a common basis with Fasanmade *et al.* 2011, Kantasiripitak *et al.* 2021 and Passot *et al.* 2016. Edlund *et al.* 2017 was included in eight of them. The mean weight and frequency of each infliximab model within the top 1000 ensembles is reported in **Table S10**. Kantasiripitak *et al.* 2021 and Passot *et al.* 2016 were the most frequently selected models, present in 79.7% and 77.3% of the top 1000 ensembles, respectively. Passot *et al.* 2016 also had the highest mean weight (15.6%), followed by Fasanmade *et al.* 2011 (13.3%). At the other end, Brandse *et al.* 2016 had a mean weight below 1% (0.3%) and Brandse *et al.* 2017 did not appear in any of the top 1000 ensembles.

### 3. Generalisability to other drugs: cefepime model averaging

#### 3.1. Method

##### 3.1.1. Literature review, data collection, model implementation, patients and data

All of these aspects for cefepime have been described in detail elsewhere [unpublished data]; briefly, a literature review identified popPK models of cefepime in critically ill adults, building on the systematic review by Hansel *et al.* 2024 ^53^, and complemented by a PubMed and Google Scholar search up to January 2025. This selection process identified 9 popPK models for cefepime: Roos *et al.* 2006, Jonckheere *et al.* 2016, Barreto *et al.* 2023a, Seo *et al.* 2023, Bilal *et al.* 2023, Carlier *et al.* 2015, Barreto *et al.* 2023b, An *et al.* 2023, and Delattre *et al.* 2012 ^54–62^.

A retrospective dataset was extracted from patient records collected between March and May 2025 at Rennes University Hospital, including 20 patients who had received at least one cefepime administration with at least two available plasma concentration measurements. Cefepime was given as a 2g loading dose over 30 minutes followed by a 6g/day maintenance dose administered by continuous infusion, with dose reduction in patients with renal impairment. Collected covariates comprised pathophysiological parameters (sex, age, weight, height, serum and urinary creatinine, urine output, cardiac index, fluid balance, end-stage renal disease, albumin, mean arterial pressure, mechanical ventilation, sepsis) and treatment-related data (dose, infusion duration, dosing frequency, treatment initiation, concomitant vasopressors, renal replacement therapy, ECMO). Lean, ideal and adjusted body weight were derived as follows :

Lean body weight (LBW) in kg ^2^ :

$$LBW_{males}=0.407\times weight \left( kg \right)+0.267\times height (cm)-19.2$$

$$LBW_{females}=0.252\times weight \left( kg \right)+0.473\times height (cm)-48.3$$

Ideal body weight (IBW) in kg ^63^ :

$$IBW=45.4+0.89 \times\left( height \left( cm \right)-152.4 \right)+4.5 (if male)$$

Adjusted body weight (ABW) in kg ^64^ :

$ABW=IBW+0.4 \times(weight (kg)-IBW)$

Renal function was estimated using the three following serum creatinine-based equations:

Estimated glomerular filtration rate (eGFR) in mL/min/1.73 m² with MDRD formula ^3^:

$${eGFR}_{MDRD}=175\times{(\frac{creat}{88.4})}^{-1.154}\times{age}^{-0.203}\times(0.742 if female)$$

Where creat is in µmol/L, and age in years.

eGFR in mL/min with Cockcroft and Gault formula ^4^*:*

$${eGFR}_{cockcroft}=1.23\times weight\times\frac{(140-age)}{creat}\times0.845 \left( if female \right)$$

Where weight is in kg, age in years and creat in µmol/L.

eGFR in mL/min/1.73 m² with CKD-EPI formula ^65^:

$${eGFR}_{CKD\_EPI}=142\times min({\frac{Scr}{k},1)}^{\alpha}\times max({\frac{Scr}{k},1)}^{-1.2}\times{0.9938}^{age} \times(1.012 if female)$$

Where Scr is in mg/dL, k is 0.7 for females and 0.9 for males, alpha is -0.241 for females and -0.302 for males, and age is in years.

No covariate value was missing. Covariate distributions are reported in **Table S8.** Plasma concentrations were measured by liquid chromatography with UV detection according to the method of Verdier *et al.* ^66^, with quantification limits of 2 and 250 µg/mL; values outside this range were excluded from the external evaluation. The project was approved by the Ethics Committee of Rennes University Hospital (approval number 26.96). This study was conducted in accordance with the principles of the Declaration of Helsinki, and, given its retrospective design and the use of previously collected, pseudonymised health data, prior written informed consent was not required. Patients were informed of the objectives of the research and of their rights through an information notice published on the transparency portal of the CHU de Rennes website.

##### 3.1.2. Evaluation design and exhaustive evaluation of all cefepime model ensembles

Models were implemented using Monolix® and Simulx® based on the information reported in the source publications. For the 9 models, concentrations at occasion 2 were computed from the individual parameters, derived from EBEs by MAP estimation using observations at occasion 1. The same weighting scheme as infliximab was used, based on $W_{m,i}^{OFV}$. For each model ensemble, RRMSE was computed.

#### 3.2. Results

The mean RRMSE per ensemble size decreased as more models were included in ensembles, with again only marginal further improvement beyond three models (from 34.3% for 3-model ensembles to 33.3% for the full 9-model ensemble), consistent with a plateau effect (**Figure S9**). Variability between ensembles of the same size also narrowed progressively with increasing size. Across all ensembles, the best-performing ensemble comprised only two models (Roos *et al.* 2006 and Bilal *et al.* 2023), with an RRMSE of 28.7% (**Table S11**). The ten best-performing ensembles included from 1 to 4 models (RRMSE 28.7-30%). Bilal *et al.* 2023 was included in all ten ensembles, while Roos *et al.* 2006 and Barreto *et al.* 2023a were each present in five of them. The mean weight and frequency of each cefepime model within the top 30 ensembles (as only 511 ensembles were possible with 9 models) is reported in **Table S12**. Bilal *et al.* 2023 was the most frequently selected model, present in all 30 top ensembles, and was the model with the highest mean weight (38%), followed by Barreto *et al.* 2023a (mean weight 21.6%).

### 4. Supplementary Figures and tables.

**Table S1**. Therapeutic range for Tacrolimus AUC.

| Renal transplant | | |
| --- | --- | --- |
| Post-Transplantation period | ${AUC}_{0-12}$ (ng.h/mL) | ${AUC}_{0-24}$ (ng.h/mL) |
| 0 – 3 months | 150 – 180 | 300 - 360 |
| 3 months – 1 year | 120 – 150 | 240 – 300 |
| > 1 year | 75 – 95 | 150 - 190 |
| Liver transplant | | |
| Post-Transplantation period | ${AUC}_{0-12}$ (ng.h/mL) | ${AUC}_{0-24}$ (ng.h/mL) |
| 0 – 3 months | 100 – 120 | 200 – 240 |
| > 3 months | 80 – 110 | 160 - 220 |

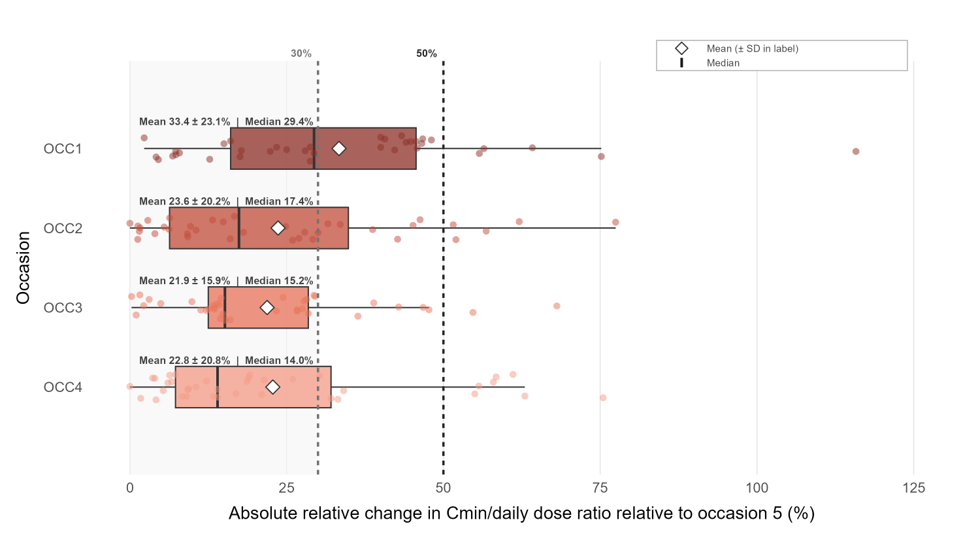
**Figure S1.** Absolute relative change in Cmin/daily dose ratio relative to occasion 5 for each previous occasion. Results show absolute median and absolute mean ± standard deviation.

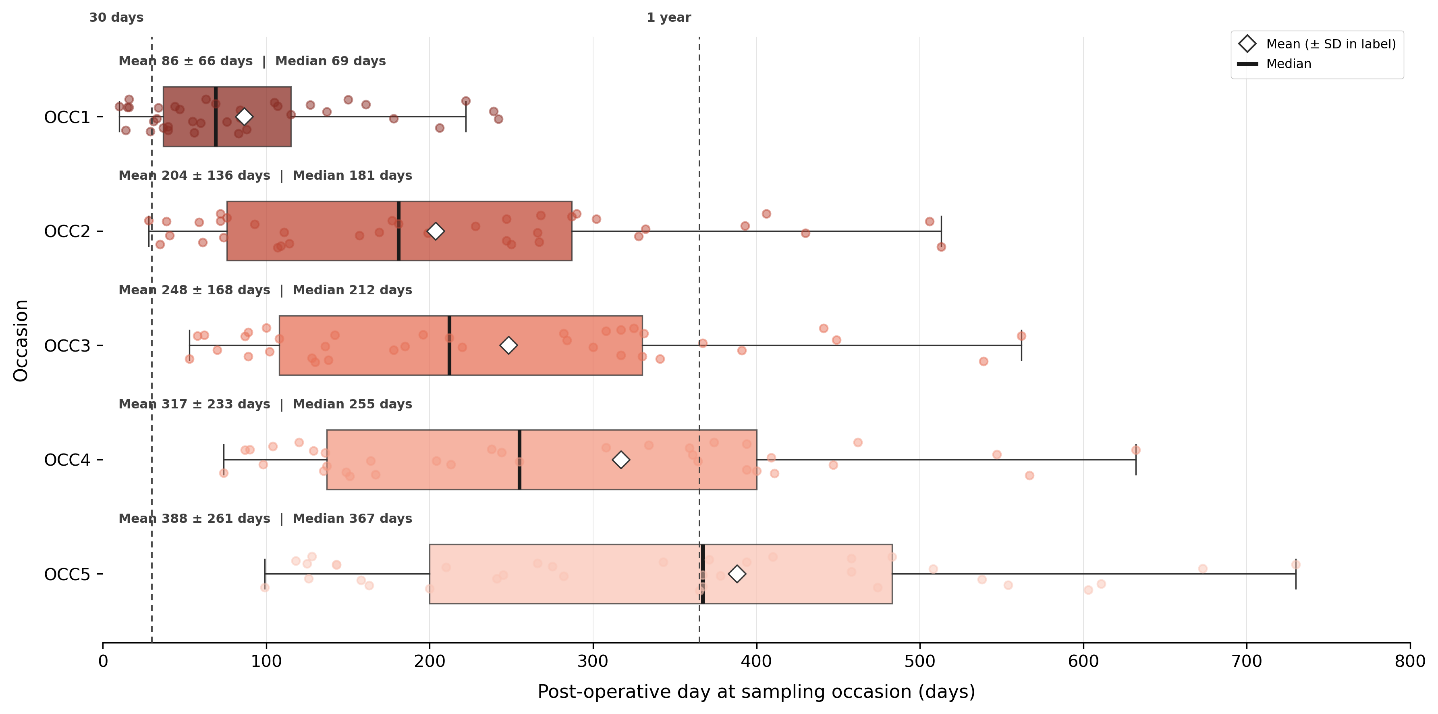
**Figure S2.** Post-operative days distribution for each occasion.

**Table S2.** Mean, minimum, and maximum weight (%) of each tacrolimus model within the top 1000 ensembles when the Cmin at occasion 5 was used for the estimation of the EBEs to predict the AUC at occasion 5, and frequency of each model among the top 1000 ensembles.

| **Model** | **Mean (%)** | **Min (%)** | **Max (%)** | **Frequency (%)** |
| --- | --- | --- | --- | --- |
| Lu 2019 | 21.9 | 0 | 61.1 | 97.3 |
| Zhu 2014 | 13 | 0 | 38.5 | 94.5 |
| Mohammed Ali 2023 | 11.3 | 0 | 61.5 | 75.3 |
| Bergmann 2014 | 5.3 | 0 | 47.3 | 27.5 |
| Martial 2021 | 5.2 | 0 | 39.5 | 30.4 |
| Moes 2016 | 5.2 | 0 | 44.6 | 25.7 |
| Ogasawara 2013 | 4.9 | 0 | 45.2 | 23.6 |
| Paschier 2023 | 4.9 | 0 | 69.1 | 25.4 |
| Grover 2011 | 4.5 | 0 | 37.5 | 27.9 |
| Sikma 2020 | 4.3 | 0 | 72.8 | 19.9 |
| Størset 2014 | 4.3 | 0 | 49.5 | 20.8 |
| Woillard 2011 | 4.2 | 0 | 47.3 | 18.8 |
| Allard 2019 | 3.4 | 0 | 35.7 | 17.1 |
| Rong 2019 | 2.1 | 0 | 35.2 | 9.5 |
| Benkali 2010 | 1.6 | 0 | 43.3 | 7.5 |
| Andreu 2015 | 1.2 | 0 | 44.1 | 5.3 |
| Kamp 2023 | 1.1 | 0 | 33.5 | 5.2 |
| Benkali 2009 | 0.4 | 0 | 3.2 | 31.7 |
| Campagne 2018 | 0.4 | 0 | 44.7 | 1.6 |
| Press 2009 | 0.3 | 0 | 36 | 1 |
| Teng 2022 | 0.3 | 0 | 10 | 3.9 |
| Andreu 2017 | 0 | 0 | 0 | 0 |
| Andrews 2019 | 0 | 0 | 0 | 0 |
| Han 2014 | 0 | 0 | 0 | 0 |
| Staatz 2003 | 0 | 0 | 0 | 0 |
| Vadcharavivad 2016 | 0 | 0 | 0 | 0 |
| Zhang 2022 | 0 | 0 | 0 | 0 |

**Figure S3.** Distribution of RRMSE across all tacrolimus model ensembles grouped by ensemble size (from 1 to a maximum of 27 models per ensemble), when the occasion 1, 2, 3, 4 observations were used for the estimation of the EBEs to predict the occasion 5 AUC. The mean, standard deviation, minimum, and maximum were computed exactly over all ensembles of a given size, whereas the scatter shows a random subsample of up to 10,000 ensembles per ensemble size.

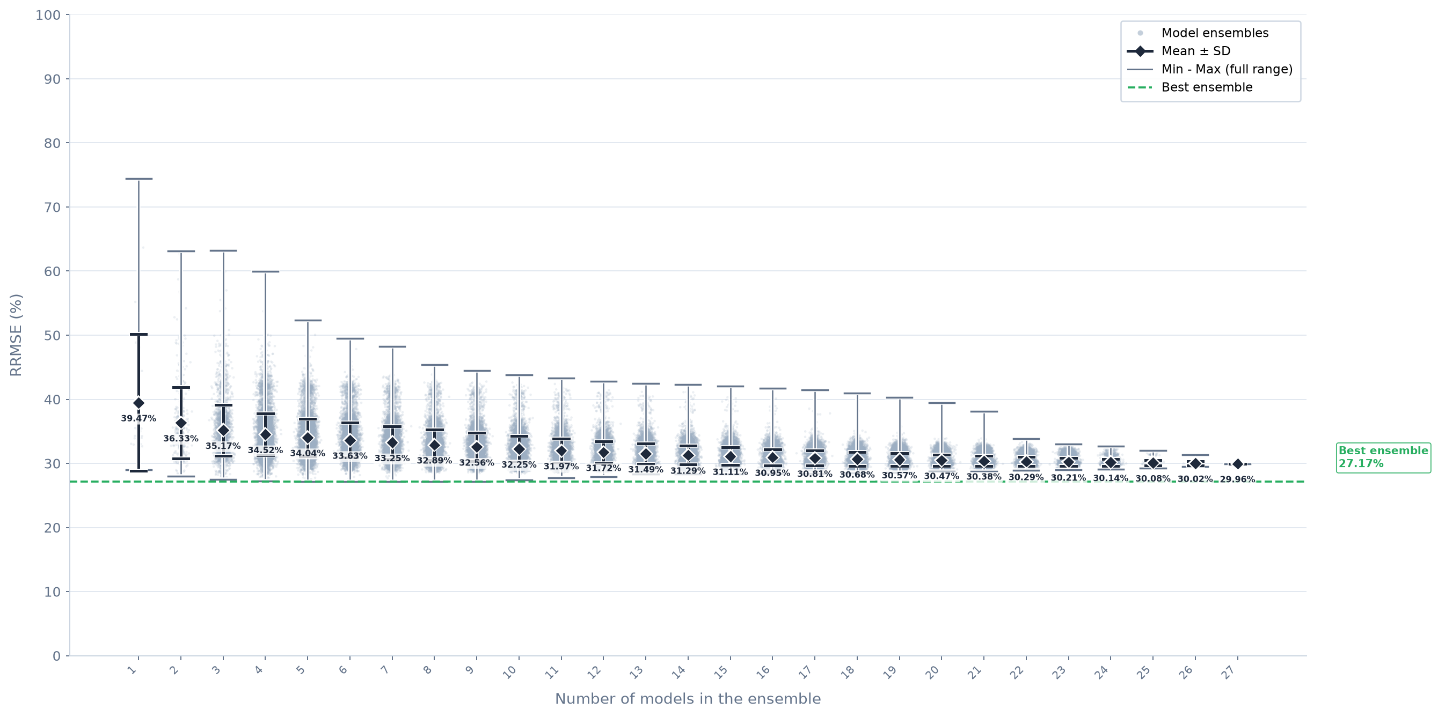

**Table S3.** Mean, minimum, and maximum weight (%) of each tacrolimus model within the top 1000 ensembles using occasion 1,2,3,4 observations to predict occasion 5 AUC, and frequency of each model among the top 1000 ensembles.

| **Model** | **Mean (%)** | **Min (%)** | **Max (%)** | **Frequency (%)** |
| --- | --- | --- | --- | --- |
| Zhu 2014 | 26.9 | 0 | 59.5 | 99.3 |
| Vadcharavivad 2016 | 20.9 | 0 | 67 | 60.5 |
| Kamp 2023 | 11 | 0 | 56.4 | 39.3 |
| Mohammed Ali 2023 | 6.5 | 0 | 28.5 | 53.1 |
| Press 2009 | 6.5 | 0 | 76.8 | 14.6 |
| Martial 2021 | 5.3 | 0 | 29.4 | 34.3 |
| Zhang 2022 | 4.8 | 0 | 44.1 | 49.1 |
| Rong 2019 | 4.7 | 0 | 15.5 | 86.5 |
| Andreu 2015 | 3.8 | 0 | 41 | 35.9 |
| Grover 2011 | 2.7 | 0 | 31.4 | 55.3 |
| Andreu 2017 | 1.7 | 0 | 5.1 | 54.7 |
| Benkali 2009 | 1.2 | 0 | 26.8 | 51.1 |
| Lu 2019 | 1.2 | 0 | 35 | 3.5 |
| Allard 2019 | 0.8 | 0 | 24.5 | 3.5 |
| Han 2014 | 0.6 | 0 | 5 | 50.6 |
| Sikma 2020 | 0.6 | 0 | 11.5 | 8.4 |
| Ogasawara 2013 | 0.4 | 0 | 13.2 | 3.5 |
| Benkali 2010 | 0.1 | 0 | 16.3 | 0.7 |
| Teng 2022 | 0 | 0 | 3 | 1.6 |
| Staatz 2003 | 0 | 0 | 2.9 | 1 |
| Campagne 2018 | 0 | 0 | 5.6 | 0.8 |
| Andrews 2019 | 0 | 0 | 0 | 0 |
| Bergmann 2014 | 0 | 0 | 0 | 0 |
| Moes 2016 | 0 | 0 | 0 | 0 |
| Paschier 2023 | 0 | 0 | 0 | 0 |
| Størset 2014 | 0 | 0 | 0 | 0 |
| Woillard 2011 | 0 | 0 | 0 | 0 |

**Figure S4.** Single model approach evaluation assessing the RRMSE for each model for the two sets of observations evaluated (Cmin at occasion 5 and observations at occasions 1,2,3 and 4 to AUC at occasion 5).

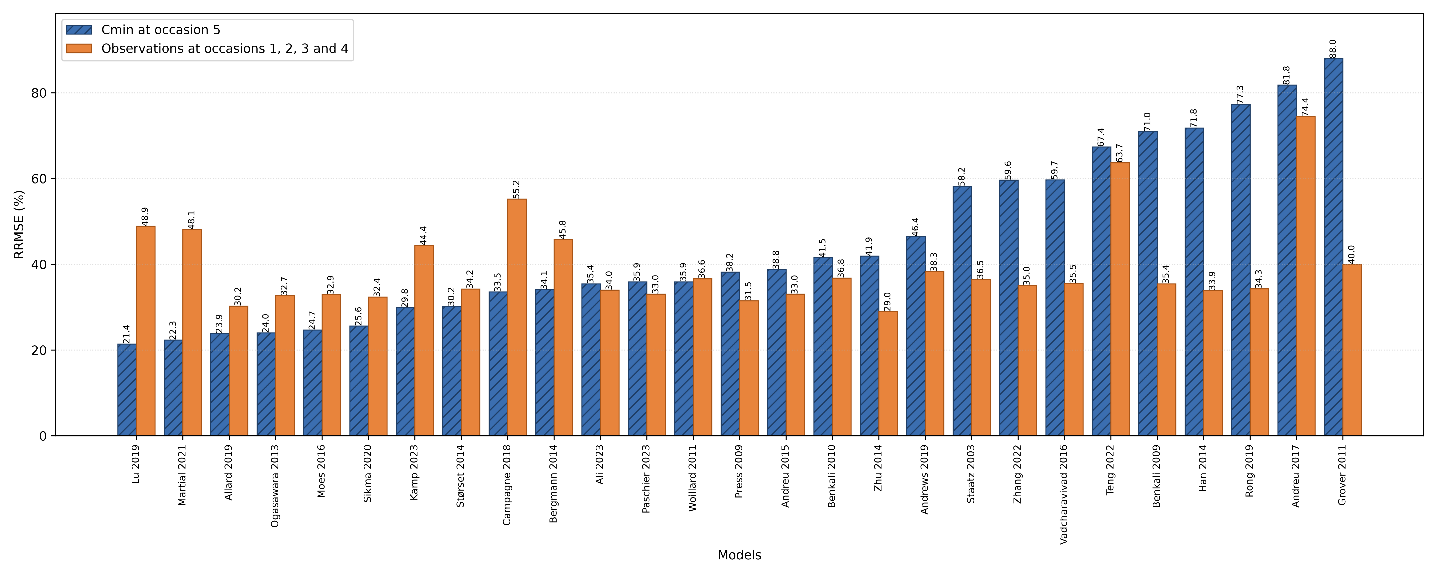

**Figure S5.** Pairwise Pearson correlation of occasion 5 AUC individual relative prediction error across the 9 tacrolimus popPK models from E_CR10. Occasion 5 AUC predicted from EBEs estimated with (A) the concentrations from occasions 1, 2, 3, 4, and (B) the Cmin at occasion 5.

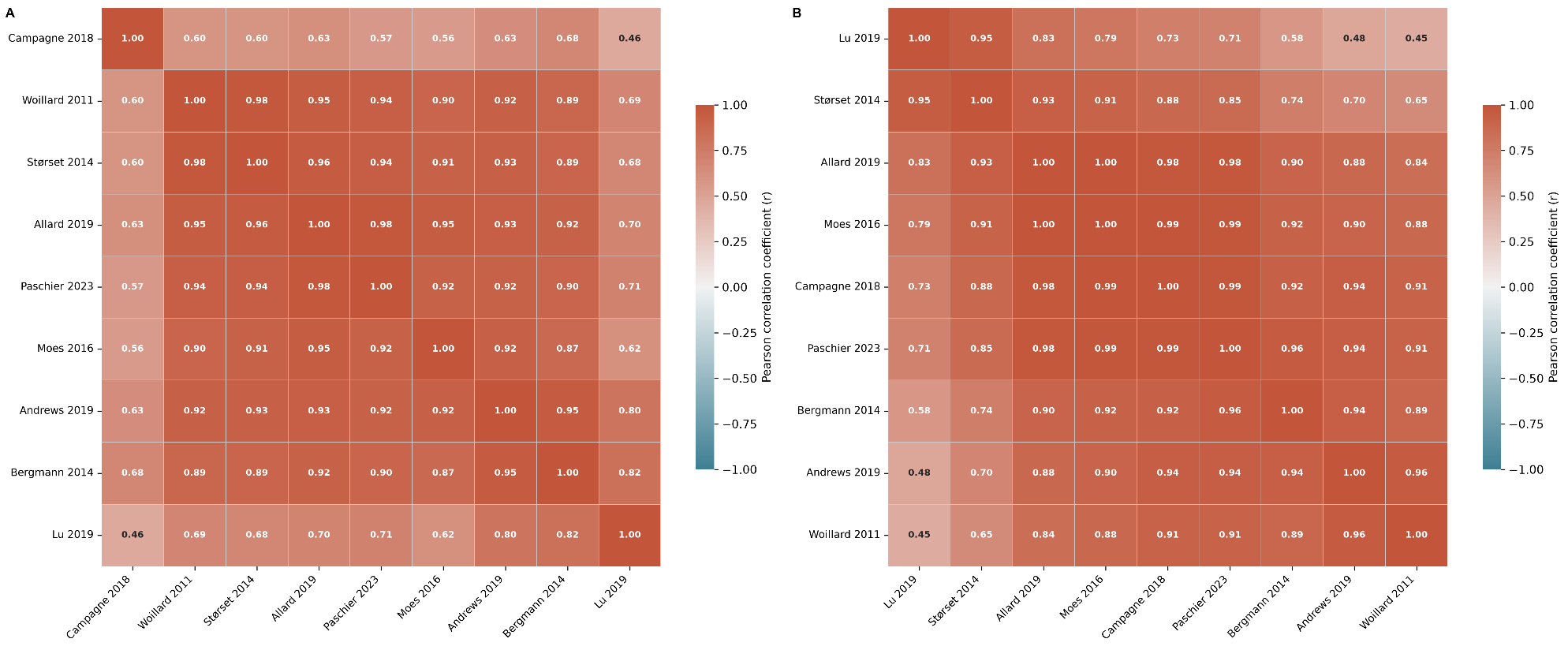

**Table S4.** RRMSE (%) of model ensembles combining a fixed base set with one interchangeable model drawn from a correlated group, for the two sets of observations used for the estimation of the EBEs (occasion 1, 2, 3, 4 concentrations and occasion 5 Cmin).

| **MAP estimation using occasion 1,2,3,4 observations** | | | | | |
| --- | --- | --- | --- | --- | --- |
| **Fixed base** | **RRMSE** | | | | |
| Andrews 2019 + Bergmann 2014 + Campagne 2018 + Lu 2019 + | Moes 2016 | Allard 2019 | Paschier 2023 | Størset 2014 | Woillard 2011 |
| RRMSE (%) | 29,15 | 30,21 | 30,87 | 34,21 | 34,39 |
| **MAP estimation using Cmin at occasion 5** | | | | | |
| **Fixed base** | **RRMSE** | | | | |
| Andrews 2019 + Bergmann 2014 + Lu 2019 + Størset 2014 + Woillard 2011 + | Allard 2019 | Moes 2016 | Paschier 2023 | Campagne 2018 |  |
| RRMSE (%) | 26,37 | 26,69 | 26,81 | 27,73 |  |

**Figure S6.** Distribution of RRMSE (%) across all tacrolimus model ensembles as a function of ensemble size, with and without models from the best-performing ensemble (Lu 2019, Grover 2011, and Zhu 2014), using Cmin at occasion 5.

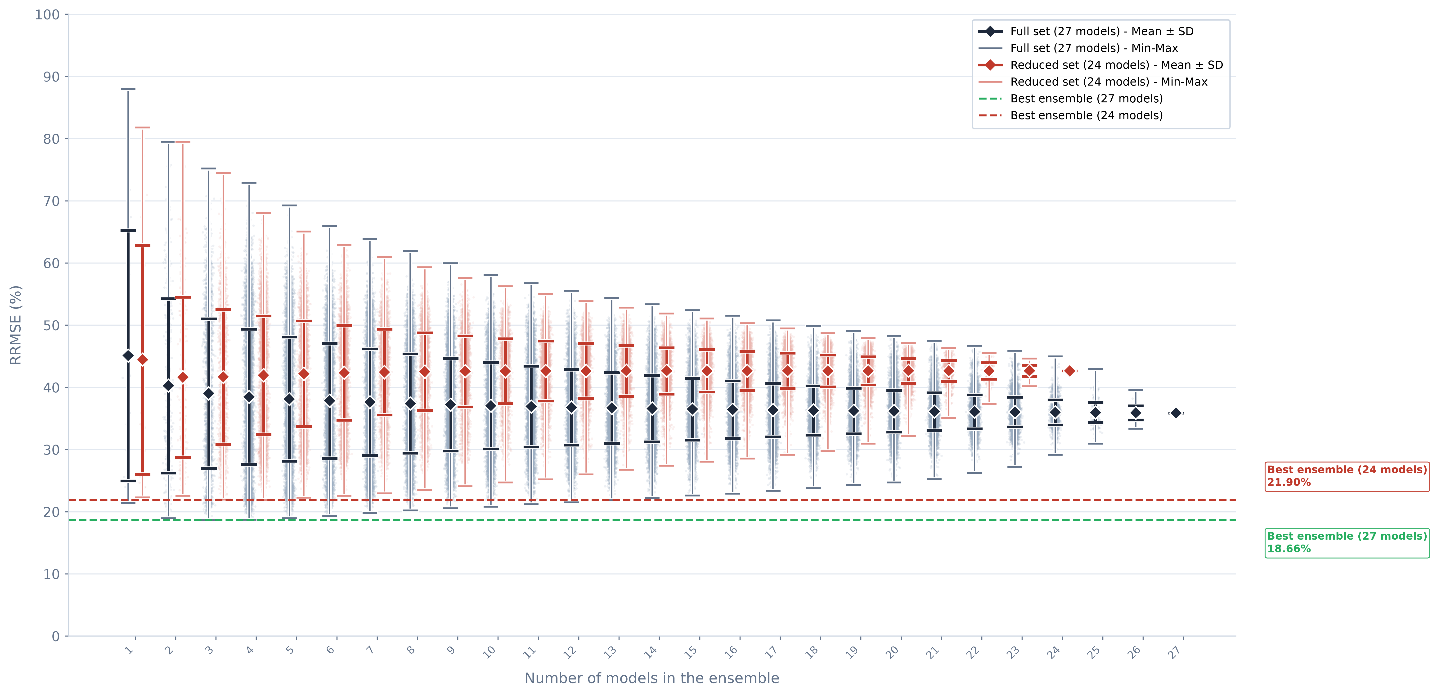

**Table S5.** Top ten of ensembles with the lowest mean training rank in the K-fold cross-validation. N models : number of models within the ensemble; RRMSE, relative root mean square error; SD, standard deviation.

| Final Rank | Models | N models | Rank  Train  mean | Rank  train_A | Rank  train_B | Rank  train_C | Rank  train_D | RRMSE  test_A | RRMSE  test_B | RRMSE  test_C | RRMSE  test_D | RRMSE  Test  mean | RRMSE  Test  SD | Rank  test |
| --- | --- | --- | --- | --- | --- | --- | --- | --- | --- | --- | --- | --- | --- | --- |
| 1 | Lu 2019 + Paschier 2023 + Mohammed Ali 2023 + Zhu 2014 | 4 | 16.25 | 8 | 6 | 6 | 45 | 19.87 | 20.06 | 12.26 | 21.21 | 18.35 | 4.1 | 1 |
| 2 | Lu 2019 + Mohammed Ali 2023 + Grover 2011 + Zhu 2014 | 4 | 18.5 | 28 | 25 | 2 | 19 | 18.54 | 19.56 | 14.52 | 21.85 | 18.62 | 3.06 | 7 |
| 3 | Lu 2019 + Andreu 2015 + Zhu 2014 | 3 | 23.25 | 41 | 7 | 8 | 37 | 18.47 | 20.89 | 13.1 | 22.3 | 18.69 | 4.05 | 8 |
| 4 | Lu 2019 + Paschier 2023 + Mohammed Ali 2023 + Benkali 2009 + Zhu 2014 | 5 | 25.75 | 17 | 29 | 14 | 43 | 19.85 | 20.16 | 12.1 | 22.09 | 18.55 | 4.41 | 5 |
| 5 | Lu 2019 + Mohammed Ali 2023 | 2 | 30.5 | 34 | 22 | 4 | 62 | 18.74 | 20.26 | 14.07 | 21.91 | 18.74 | 3.37 | 14 |
| 6 | Lu 2019 + Mohammed Ali 2023 + Grover 2011 + Martial 2021 + Zhu 2014 | 5 | 34.75 | 53 | 33 | 10 | 43 | 18.61 | 20.71 | 13.28 | 22.62 | 18.81 | 4.03 | 18 |
| 7 | Lu 2019 + Størset 2014 + Zhu 2014 | 3 | 39.5 | 20 | 33 | 3 | 102 | 19.51 | 19.99 | 13.89 | 21.39 | 18.69 | 3.3 | 9 |
| 8 | Lu 2019 + Mohammed Ali 2023 + Benkali 2009 + Grover 2011 + Zhu 2014 | 5 | 46.25 | 42 | 121 | 4 | 18 | 18.51 | 19.58 | 14.35 | 22.74 | 18.8 | 3.46 | 17 |
| 9 | Lu 2019 + Grover 2011 + Zhu 2014 | 3 | 47.5 | 9 | 175 | 1 | 5 | 19.46 | 17.89 | 14.27 | 22.06 | 18.42 | 3.26 | 3 |
| 10 | Lu 2019 + Størset 2014 + Mohammed Ali 2023 + Zhu 2014 | 4 | 51.25 | 46 | 22 | 7 | 130 | 18.77 | 20.82 | 14.04 | 21.89 | 18.88 | 3.48 | 22 |

**Table S6.** Optimal model ensemble selected on the mean training rank and ensembles ranked first within each individual fold. RRMSE, relative root mean square error.

| Status | Model ensemble | N  Models | Mean training rank | Rank  train_A | Rank  train_B | Rank  train_C | Rank  train_D | RRMSE  test_A | RRMSE  test_B | RRMSE  test_C | RRMSE  test_D | RRMSE  Test  mean | RRMSE  Test  sd | Rank  test |
| --- | --- | --- | --- | --- | --- | --- | --- | --- | --- | --- | --- | --- | --- | --- |
| Overall  Best ensemble | Lu 2019 + Paschier 2023 + Mohammed Ali 2023 + Zhu 2014 | 4 | 16.25 | 8 | 6 | 6 | 45 | 19.87 | 20.06 | 12.26 | 21.21 | 18.35 | 4.1 | 1 |
| Best in fold A | Mohammed Ali 2023 + Zhu 2014 | 2 | 4732.75 | 1 | 6311 | 59 | 12560 | 25.63 | 17.76 | 11.79 | 18.49 | 18.42 | 5.67 | 2 |
| Best in fold B | Lu 2019 + Paschier 2023 | 2 | 266 | 1041 | 1 | 12 | 10 | 14.07 | 23.28 | 14.05 | 23.76 | 18.79 | 5.47 | 16 |
| Best in fold C | Lu 2019 + Grover 2011 + Zhu 2014 | 3 | 47.5 | 9 | 175 | 1 | 5 | 19.46 | 17.89 | 14.27 | 22.06 | 18.42 | 3.26 | 3 |
| Best in fold D | Lu 2019 + Woillard 2011 + Mohammed Ali 2023 | 3 | 466.75 | 1824 | 2 | 40 | 1 | 13.04 | 23.26 | 12.95 | 25.18 | 18.61 | 6.53 | 6 |

**Table S7.** Patient and dosing characteristics. Infliximab PK database (stratified by occasion).

| **Characteristic** | **OCC 1 (n=187)** | **OCC 2 (n=187)** | **OCC 3 (n=187)** | **OCC 4 (n=187)** | **OCC 5 (n=187)** | **Overall (n=187)** |
| --- | --- | --- | --- | --- | --- | --- |
| Study design |  |  |  |  |  |  |
| N patients | 187 | 187 | 187 | 187 | 187 | 187 |
| N observations | 187 | 187 | 187 | 187 | 187 | 935 |
| Infliximab concentration (mg/L) |  |  |  |  |  |  |
| Median | 4.20 | 4.60 | 4.40 | 4.50 | 5 | 4.50 |
| Min | 0.10 | 0.10 | 0.10 | 0.10 | 1 | 0.10 |
| Max | 56.90 | 44.30 | 47.80 | 43.60 | 48.40 | 56.90 |
| [Q1–Q3] | 2.80–7.95 | 3.05–7.40 | 3.0–6.65 | 2.95–7.45 | 3.30–7.55 | 3.0–7.40 |
| Dose |  |  |  |  |  |  |
| Median dose (mg) | 370 (210–1340) | 370 (220–1530) | 385 (215–1500) | 400 (210–1530) | 487.5 (210–1530) | 390 (210–1530) |
| Median dose/kg (mg/kg) | 5 (4.20–10) | 5 (4.20–10) | 5 (4.20–10) | 5 (4.20–10) | 5 (5–10) | 5 (4.20–10) |
| Continuous covariates - Median (min-max) |  |  |  |  |  |  |
| Age (years) | 36 (18–96) | 36 (18–96) | 36 (18–96) | 36 (18–96) | 37 (19–97) | 36 (18–97) |
| Weight (kg) | 68 (42–150) | 68.5 (41–152) | 68 (42–150) | 69 (42–153) | 69 (42–153) | 68 (41–153) |
| Albumin (g/L) | 43 (24.8–50.6) | 43.2 (24.8–50.6) | 43 (24.8–52.5) | 43 (24.8–52.5) | 43 (24.8–52.5) | 43 (24.8–52.5) |
| CRP (mg/L) | 6 (0.2–586) | 5 (0.2–404) | 4 (0.3–432) | 5 (0.3–124) | 4 (0.2–128) | 5 (0.2–586) |
| Faecal calprotectin (µg/g) | 0 (0–1255) | 0 (0–800) | 0 (0–314) | 0 (0–503) | 0 (0–800) | 0 (0–1255) |
| Harvey-Bradshaw Index | 0 (0–22) | 0 (0–11) | 0 (0–11) | 0 (0–16) | 0 (0–17) | 0 (0–22) |
| PDAI score | 0 (0–5) | 0 (0–10) | 0 (0–9) | 0 (0–6) | 0 (0–9) | 0 (0–10) |
| Categorical covariates - N (%) |  |  |  |  |  |  |
| Sex |  |  |  |  |  |  |
| Male | 90 (48%) | 90 (48%) | 90 (48%) | 90 (48%) | 90 (48%) | 90 (48%) |
| Female | 97 (52%) | 97 (52%) | 97 (52%) | 97 (52%) | 97 (52%) | 97 (52%) |
| Anti-drug antibodies |  |  |  |  |  |  |
| No | 187 (100%) | 187 (100%) | 187 (100%) | 187 (100%) | 187 (100%) | - |
| Maintenance therapy |  |  |  |  |  |  |
| Initiation | 42 (22%) | 5 (3%) | 1 (1%) | 0 (0%) | 0 (0%) | - |
| Maintenance | 145 (78%) | 182 (97%) | 186 (99%) | 187 (100%) | 187 (100%) | - |
| Co-treatment |  |  |  |  |  |  |
| No | 115 (61%) | 117 (63%) | 122 (65%) | 121 (65%) | 124 (66%) | - |
| Yes | 72 (39%) | 70 (37%) | 65 (35%) | 66 (35%) | 63 (34%) | - |
| Immunosuppressant (any) |  |  |  |  |  |  |
| No | 119 (64%) | 119 (64%) | 124 (66%) | 123 (66%) | 126 (67%) | - |
| Yes | 68 (36%) | 68 (36%) | 63 (34%) | 64 (34%) | 61 (33%) | - |

**Figure S7.** Prisma flow chart for infliximab systematic literature review.

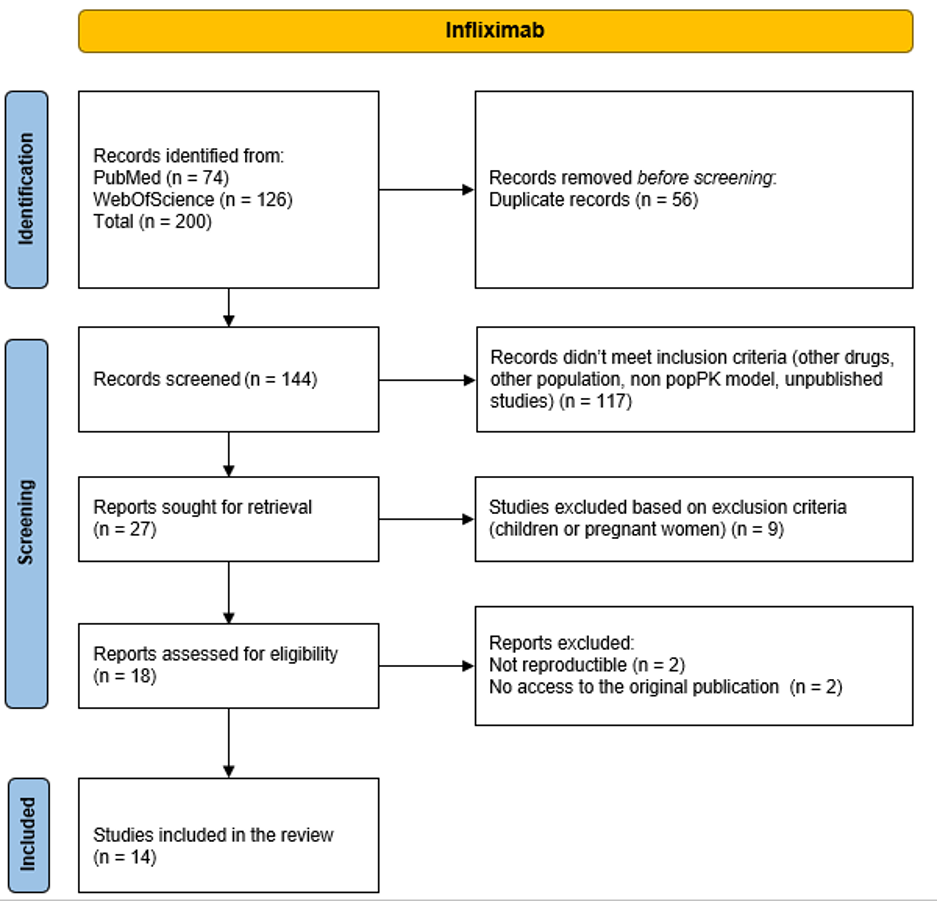

**Table S8.** Patient characteristics and covariate distribution for cefepime dataset.

| **Characteristic** | **OCC 1 (n=20)** | **OCC 2 (n=20)** | **Overall (n=20)** |
| --- | --- | --- | --- |
| Study design |  |  |  |
| N patients | 20 | 20 | 20 |
| N observations | 20 | 20 | 40 |
| Cefepime concentration (mg/L) |  |  |  |
| Median | 55.40 | 39.45 | 42.80 |
| Min | 13.20 | 11.90 | 11.90 |
| Max | 163.30 | 97.50 | 163.30 |
| Q1–Q3 | 37.33–63.45 | 29.23–47.70 | 32.75–57.33 |
| Continuous covariates - Median (min-max) |  |  |  |
| Age (years) | 65 (36–74) | 65 (36–74) | 65 (36–74) |
| Height (cm) | 172 (154–188) | 172 (154–188) | 172 (154–188) |
| Weight (kg) | 85.8 (54–120) | 81.8 (52.5–122) | 83.2 (52.5–122) |
| Lean body weight (kg) | 58.1 (44.3–72.1) | 58.2 (44.3–72.1) | 58.2 (44.3–72.1) |
| Body surface area (m²) | 2 (1.5–2.4) | 2 (1.5–2.4) | 2 (1.5–2.4) |
| Serum creatinine (µmol/L) | 77 (46–335) | 66 (45–364) | 70 (45–364) |
| Urine creatinine (mmol/L) | 3.5 (0–7.4) | 2.1 (0–6.7) | 2.5 (0–7.4) |
| Cockcroft – Gault (mL/min) | 105.1 (19.7–178.6) | 101.9 (17.8–231.8) | 102.1 (17.8–231.8) |
| CKD-EPI (mL/min/1.73m²) | 83.8 (15.3–121.2) | 94.7 (13.8–121.2) | 87.1 (13.8–121.2) |
| Mean arterial pressure (mmHg) | 83 (61–111) | 84 (59–123) | 83 (59–123) |
| Categorical covariates - N (%) |  |  |  |
| Sex |  |  |  |
| Female | 6 (30%) | 6 (30%) | 6 (30%) |
| Male | 14 (70%) | 14 (70%) | 14 (70%) |
| End-stage renal disease |  |  |  |
| No | 20 (100%) | 20 (100%) | - |
| ECMO |  |  |  |
| No | 18 (90%) | 19 (95%) | - |
| Yes | 2 (10%) | 1 (5%) | - |
| Dialysis |  |  |  |
| No | 18 (90%) | 17 (85%) | - |
| Yes | 2 (10%) | 3 (15%) | - |
| Mechanical ventilation |  |  |  |
| No | 3 (15%) | 2 (10%) | - |
| Yes | 15 (75%) | 15 (75%) | - |
| Partial | 2 (10%) | 3 (15%) | - |
| Sepsis |  |  |  |
| No | 19 (95%) | 19 (95%) | - |
| Yes | 1 (5%) | 1 (5%) |  |

**Figure S8.** Distribution of RRMSE across all model ensembles grouped by ensemble size (from 1 to a maximum of 14 models per ensemble), for infliximab use case.

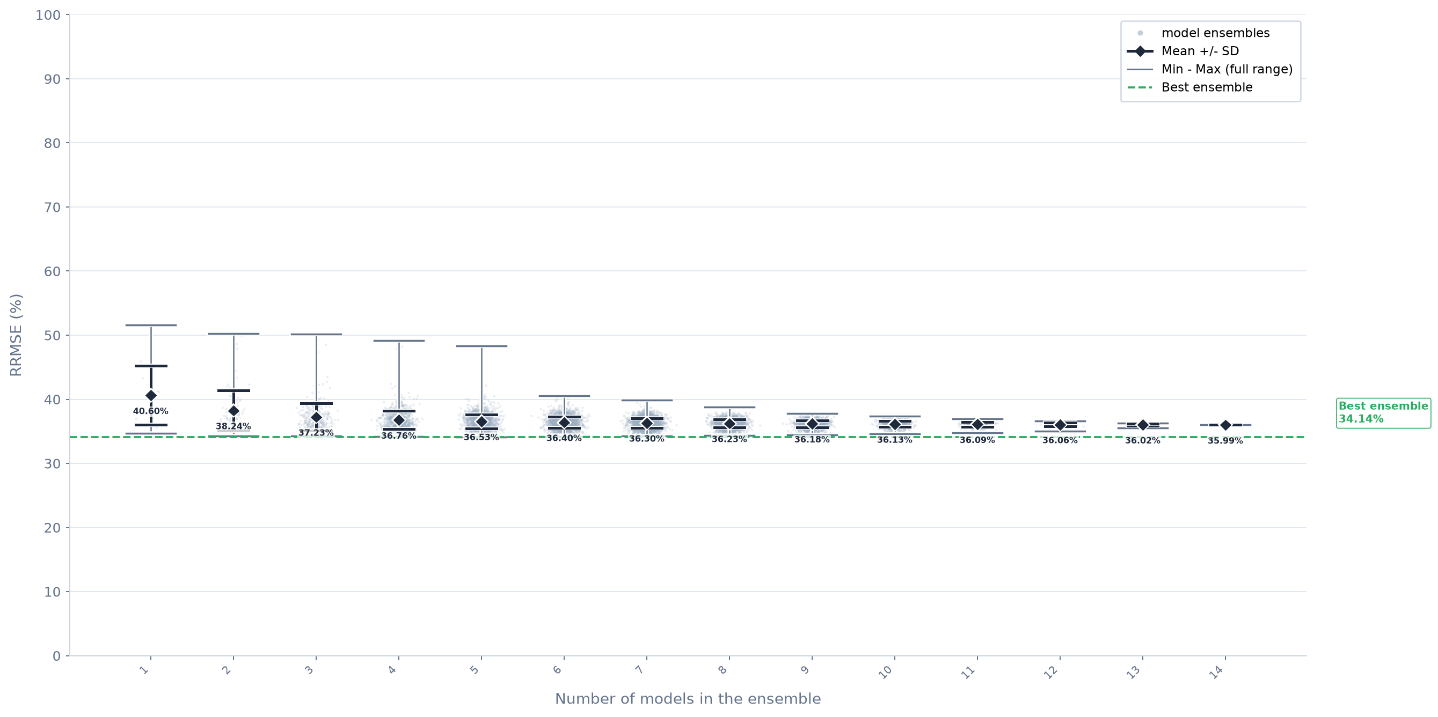

**Table S9.** Top ten ensembles with the lowest RRMSE, identified through exhaustive evaluation of infliximab model-averaging ensembles. N models : number of models within the ensemble; RRMSE, relative root mean square error.

| **Model ensemble** | **N models** | **RRMSE** | **rank** |
| --- | --- | --- | --- |
| Edlund 2017 + Fasanmade 2011 + Kantasiripitak 2021 + Magro 2024 + Passot 2016 | 5 | 34.14 | 1 |
| Edlund 2017 + Fasanmade 2011 + Kantasiripitak 2021 + Passot 2016 | 4 | 34.16 | 2 |
| Buurman 2015 + Dreesen 2021 + Edlund 2017 + Fasanmade 2011 + Kantasiripitak 2021 + Passot 2016 | 6 | 34.21 | 3 |
| Buurman 2015 + Edlund 2017 + Fasanmade 2011 + Kantasiripitak 2021 + Passot 2016 | 5 | 34.21 | 3 |
| Buurman 2015 + Dotan 2014 + Edlund 2017 + Fasanmade 2011 + Kantasiripitak 2021 + Passot 2016 | 6 | 34.23 | 5 |
| Fasanmade 2011 + Kantasiripitak 2021 + Passot 2016 | 3 | 34.25 | 6 |
| Fasanmade 2011 + Kantasiripitak 2021 + Magro 2024 + Passot 2016 | 4 | 34.25 | 6 |
| Buurman 2015 + Dotan 2014 + Dreesen 2021 + Edlund 2017 + Fasanmade 2011 + Kantasiripitak 2021 + Passot 2016 | 7 | 34.26 | 8 |
| Edlund 2017 + Fasanmade 2009 + Fasanmade 2011 + Kantasiripitak 2021 + Magro 2024 + Passot 2016 | 6 | 34.26 | 8 |
| Dreesen 2021 + Edlund 2017 + Fasanmade 2011 + Kantasiripitak 2021 + Magro 2024 + Passot 2016 | 6 | 34.28 | 10 |

**Table S10.** Mean, minimum, and maximum weight (%) of each infliximab model within the top 1000 ensembles for OCC5 Cmin prediction based on OCC1,2,3,4 Cmin, and frequency of each model among the top 1000 ensembles.

| **Model** | **Mean (%)** | **Min (%)** | **Max (%)** | **Frequency (%)** |
| --- | --- | --- | --- | --- |
| Passot 2016 | 15.6 | 0 | 76.9 | 77.3 |
| Fasanmade 2011 | 13.3 | 0 | 100 | 73 |
| Buurman 2015 | 9.5 | 0 | 29.7 | 64.4 |
| Kantasiripitak 2021 | 9.4 | 0 | 36 | 79.7 |
| Edlund 2017 | 9 | 0 | 22.2 | 67.8 |
| Fasanmade 2009 | 8.8 | 0 | 41.4 | 53.2 |
| Grisic 2021 | 7.3 | 0 | 42.9 | 38 |
| Berends 2019 | 6.8 | 0 | 46 | 31.2 |
| Dreesen 2021 | 6.8 | 0 | 28.7 | 46.5 |
| Dotan 2014 | 4.8 | 0 | 28.3 | 45 |
| Magro 2024 | 4.8 | 0 | 54.9 | 58.7 |
| Ternant 2018 | 3.6 | 0 | 27.1 | 25.2 |
| Brandse 2016 | 0.3 | 0 | 8.6 | 4.8 |
| Brandse 2017 | 0 | 0 | 0 | 0 |

**Figure S9.** Distribution of RRMSE across all model ensembles grouped by ensemble size (from 1 to a maximum of 9 models per ensemble), for cefepime use case.

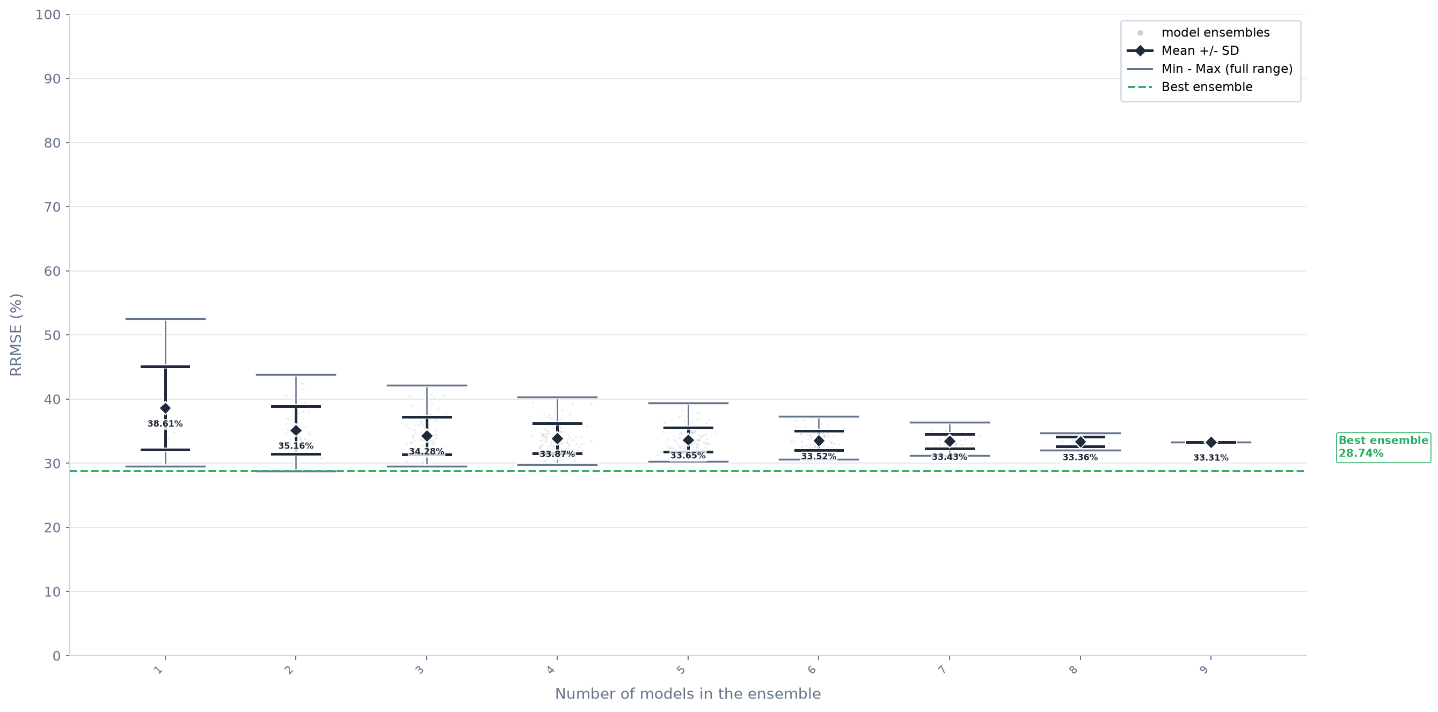

**Table S11.** Top ten ensembles with the lowest RRMSE, identified through exhaustive evaluation of cefepime model-averaging ensembles. N models : number of models within the ensemble; RRMSE, relative root mean square error.

| **Model ensemble** | **N models** | **RRMSE** | **rank** |
| --- | --- | --- | --- |
| Roos 2006 + Bilal 2023 | 2 | 28.74 | 1 |
| Jonckheere 2016 + Bilal 2023 | 2 | 29.08 | 2 |
| Bilal 2023 | 1 | 29.48 | 3 |
| Barreto 2023a + Bilal 2023 | 2 | 29.51 | 4 |
| Roos 2006 + Jonckheere 2016 + Bilal 2023 | 3 | 29.56 | 5 |
| Roos 2006 + Barreto 2023a + Bilal 2023 | 3 | 29.59 | 6 |
| Roos 2006 + Bilal 2023 + Barreto 2023b | 3 | 29.78 | 7 |
| Roos 2006 + Barreto 2023a + Bilal 2023 + Barreto 2023b | 4 | 29.81 | 8 |
| Jonckheere 2016 + Barreto 2023a + Bilal 2023 | 3 | 29.84 | 9 |
| Barreto 2023a + Bilal 2023 + Barreto 2023b | 3 | 29.9 | 10 |

**Table S12.** Mean, minimum, and maximum weight (%) of each cefepime model within top 30 ensembles for OCC2 concentrations prediction based on OCC1 concentrations, and frequency of each model among top 30 ensembles.

| **Model** | **Mean (%)** | **Min (%)** | **Max (%)** | **Frequency (%)** |
| --- | --- | --- | --- | --- |
| Bilal 2023 | 38 | 19 | 100 | 100 |
| Barreto 2023a | 21.6 | 0 | 52.8 | 66.7 |
| Roos 2006 | 11.5 | 0 | 43.2 | 46.7 |
| Barreto 2023b | 10.8 | 0 | 42 | 46.7 |
| Jonckheere 2016 | 10.7 | 0 | 39.8 | 50 |
| Seo 2023 | 3.9 | 0 | 23.1 | 36.7 |
| An 2023 | 3.5 | 0 | 28.5 | 13.3 |
| Carlier 2015 | 0 | 0 | 0 | 0 |
| Delattre 2012 | 0 | 0 | 0 | 0 |

### 5. References

1.	Mosteller RD. Simplified calculation of body-surface area. *N Engl J Med*. 1987;317(17):1098. doi:10.1056/NEJM198710223171717

2.	Boer P. Estimated lean body mass as an index for normalization of body fluid volumes in humans. *Am J Physiol*. 1984;247(4 Pt 2):F632-636. doi:10.1152/ajprenal.1984.247.4.F632

3.	Levey AS, Coresh J, Greene T, et al. Using Standardized Serum Creatinine Values in the Modification of Diet in Renal Disease Study Equation for Estimating Glomerular Filtration Rate. *Ann Intern Med*. 2006;145(4):247-254. doi:10.7326/0003-4819-145-4-200608150-00004

4.	Cockcroft DW, Gault MH. Prediction of creatinine clearance from serum creatinine. *Nephron*. 1976;16(1):31-41. doi:10.1159/000180580

5.	Allard M, Puszkiel A, Conti F, et al. Pharmacokinetics and Pharmacodynamics of Once-daily Prolonged-release Tacrolimus in Liver Transplant Recipients. *Clin Ther*. 2019;41(5):882-896.e3. doi:10.1016/j.clinthera.2019.03.006

6.	Andreu F, Colom H, Grinyó JM, Torras J, Cruzado JM, Lloberas N. Development of a population PK model of tacrolimus for adaptive dosage control in stable kidney transplant patients. *Ther Drug Monit*. 2015;37(2):246-255. doi:10.1097/FTD.0000000000000134

7.	Andreu F, Colom H, Elens L, et al. A New CYP3A5*3 and CYP3A4*22 Cluster Influencing Tacrolimus Target Concentrations: A Population Approach. *Clin Pharmacokinet*. 2017;56(8):963-975. doi:10.1007/s40262-016-0491-3

8.	Andrews LM, Hesselink DA, van Schaik RHN, et al. A population pharmacokinetic model to predict the individual starting dose of tacrolimus in adult renal transplant recipients. *Br J Clin Pharmacol*. 2019;85(3):601-615. doi:10.1111/bcp.13838

9.	Benkali K, Prémaud A, Picard N, et al. Tacrolimus population pharmacokinetic-pharmacogenetic analysis and Bayesian estimation in renal transplant recipients. *Clin Pharmacokinet*. 2009;48(12):805-816. doi:10.2165/11318080-000000000-00000

10.	Benkali K, Rostaing L, Premaud A, et al. Population pharmacokinetics and Bayesian estimation of tacrolimus exposure in renal transplant recipients on a new once-daily formulation. *Clin Pharmacokinet*. 2010;49(10):683-692. doi:10.2165/11535950-000000000-00000

11.	Bergmann TK, Hennig S, Barraclough KA, Isbel NM, Staatz CE. Population pharmacokinetics of tacrolimus in adult kidney transplant patients: impact of CYP3A5 genotype on starting dose. *Ther Drug Monit*. 2014;36(1):62-70. doi:10.1097/FTD.0b013e31829f1ab8

12.	Campagne O, Mager DE, Brazeau D, Venuto RC, Tornatore KM. Tacrolimus Population Pharmacokinetics and Multiple CYP3A5 Genotypes in Black and White Renal Transplant Recipients. *J Clin Pharmacol*. 2018;58(9):1184-1195. doi:10.1002/jcph.1118

13.	Grover A, Frassetto LA, Benet LZ, Chakkera HA. Pharmacokinetic differences corroborate observed low tacrolimus dosage in Native American renal transplant patients. *Drug Metab Dispos*. 2011;39(11):2017-2019. doi:10.1124/dmd.111.041350

14.	Han N, Ha S, Yun H yeol, et al. Population pharmacokinetic-pharmacogenetic model of tacrolimus in the early period after kidney transplantation. *Basic Clin Pharmacol Toxicol*. 2014;114(5):400-406. doi:10.1111/bcpt.12176

15.	Henin E, Govoni M, Cella M, Laveille C, Piotti G. Therapeutic Drug Monitoring Strategies for Envarsus in De Novo Kidney Transplant Patients Using Population Modelling and Simulations. *Adv Ther*. 2021;38(10):5317-5332. doi:10.1007/s12325-021-01905-5

16.	Kamp J, Zwart TC, Meziyerh S, et al. Meltdose Tacrolimus Population Pharmacokinetics and Limited Sampling Strategy Evaluation in Elderly Kidney Transplant Recipients. *Pharmaceutics*. 2023;16(1):17. doi:10.3390/pharmaceutics16010017

17.	Lu Z, Bonate P, Keirns J. Population pharmacokinetics of immediate- and prolonged-release tacrolimus formulations in liver, kidney and heart transplant recipients. *Br J Clin Pharmacol*. 2019;85(8):1692-1703. doi:10.1111/bcp.13952

18.	Martial LC, Biewenga M, Ruijter BN, et al. Population pharmacokinetics and genetics of oral meltdose tacrolimus (Envarsus) in stable adult liver transplant recipients. *Br J Clin Pharmacol*. 2021;87(11):4262-4272. doi:10.1111/bcp.14842

19.	Moes DJ a. R, van der Bent S a. S, Swen JJ, et al. Population pharmacokinetics and pharmacogenetics of once daily tacrolimus formulation in stable liver transplant recipients. *Eur J Clin Pharmacol*. 2016;72(2):163-174. doi:10.1007/s00228-015-1963-3

20.	Mohammed Ali Z, Meertens M, Fernández B, et al. CYP3A5*3 and CYP3A4*22 Cluster Polymorphism Effects on LCP-Tac Tacrolimus Exposure: Population Pharmacokinetic Approach. *Pharmaceutics*. 2023;15(12):2699. doi:10.3390/pharmaceutics15122699

21.	Ogasawara K, Chitnis SD, Gohh RY, Christians U, Akhlaghi F. Multidrug resistance-associated protein 2 (MRP2/ABCC2) haplotypes significantly affect the pharmacokinetics of tacrolimus in kidney transplant recipients. *Clin Pharmacokinet*. 2013;52(9):751-762. doi:10.1007/s40262-013-0069-2

22.	Paschier A, Destere A, Monchaud C, Labriffe M, Marquet P, Woillard JB. Tacrolimus population pharmacokinetics in adult heart transplant patients. *Br J Clin Pharmacol*. 2023;89(12):3584-3595. doi:10.1111/bcp.15857

23.	Press RR, Ploeger BA, den Hartigh J, et al. Explaining variability in tacrolimus pharmacokinetics to optimize early exposure in adult kidney transplant recipients. *Ther Drug Monit*. 2009;31(2):187-197. doi:10.1097/FTD.0b013e31819c3d6d

24.	Rong Y, Mayo P, Ensom MHH, Kiang TKL. Population Pharmacokinetic Analysis of Immediate-Release Oral Tacrolimus Co-administered with Mycophenolate Mofetil in Corticosteroid-Free Adult Kidney Transplant Recipients. *Eur J Drug Metab Pharmacokinet*. 2019;44(3):409-422. doi:10.1007/s13318-018-0525-3

25.	Sikma MA, Van Maarseveen EM, Hunault CC, et al. Unbound Plasma, Total Plasma, and Whole-Blood Tacrolimus Pharmacokinetics Early After Thoracic Organ Transplantation. *Clin Pharmacokinet*. 2020;59(6):771-780. doi:10.1007/s40262-019-00854-1

26.	Staatz CE, Willis C, Taylor PJ, Lynch SV, Tett SE. Toward better outcomes with tacrolimus therapy: population pharmacokinetics and individualized dosage prediction in adult liver transplantation. *Liver Transpl*. 2003;9(2):130-137. doi:10.1053/jlts.2003.50023

27.	Størset E, Holford N, Hennig S, et al. Improved prediction of tacrolimus concentrations early after kidney transplantation using theory-based pharmacokinetic modelling. *Br J Clin Pharmacol*. 2014;78(3):509-523. doi:10.1111/bcp.12361

28.	Teng F, Zhang W, Wang W, et al. Population pharmacokinetics of tacrolimus in Chinese adult liver transplant patients. *Biopharm Drug Dispos*. 2022;43(2):76-85. doi:10.1002/bdd.2311

29.	Vadcharavivad S, Praisuwan S, Techawathanawanna N, Treyaprasert W, Avihingsanon Y. Population pharmacokinetics of tacrolimus in Thai kidney transplant patients: comparison with similar data from other populations. *J Clin Pharm Ther*. 2016;41(3):310-328. doi:10.1111/jcpt.12396

30.	Woillard JB, de Winter BCM, Kamar N, Marquet P, Rostaing L, Rousseau A. Population pharmacokinetic model and Bayesian estimator for two tacrolimus formulations--twice daily Prograf and once daily Advagraf. *Br J Clin Pharmacol*. 2011;71(3):391-402. doi:10.1111/j.1365-2125.2010.03837.x

31.	Zhang SF, Tang BH, Wei AH, Du Y, Guan ZW, Li Y. Effect of drug combination on tacrolimus target dose in renal transplant patients with different CYP3A5 genotypes. *Xenobiotica*. 2022;52(3):312-321. doi:10.1080/00498254.2022.2064252

32.	Zhu L, Wang H, Sun X, et al. The Population Pharmacokinetic Models of Tacrolimus in Chinese Adult Liver Transplantation Patients. *J Pharm (Cairo)*. 2014;2014:713650. doi:10.1155/2014/713650

33.	Buurman DJ, Maurer JM, Keizer RJ, Kosterink JGW, Dijkstra G. Population pharmacokinetics of infliximab in patients with inflammatory bowel disease: potential implications for dosing in clinical practice. *Aliment Pharmacol Ther*. 2015;42(5):529-539. doi:10.1111/apt.13299

34.	Tron C, Lemaitre F, Bros P, et al. Quantification of infliximab and adalimumab in human plasma by a liquid chromatography tandem mass spectrometry kit and comparison with two ELISA methods. *Bioanalysis*. 2022;14(11):831-844. doi:10.4155/bio-2022-0057

35.	Franck B, Tron C, Verdier MC, et al. One Concentration Does Not Fit All: It Is Time to Personalize the Therapeutic Range of Infliximab in Crohn Disease. *Ther Drug Monit*. 2025;47(2):265-273. doi:10.1097/FTD.0000000000001251

36.	PRISMA 2020 explanation and elaboration: updated guidance and exemplars for reporting systematic reviews | The BMJ. Accessed November 27, 2024. https://www.bmj.com/content/372/bmj.n160

37.	McGowan J, Sampson M, Salzwedel DM, Cogo E, Foerster V, Lefebvre C. PRESS Peer Review of Electronic Search Strategies: 2015 Guideline Statement. *J Clin Epidemiol*. 2016;75:40-46. doi:10.1016/j.jclinepi.2016.01.021

38.	Berends SE, van Steeg TJ, Ahsman MJ, et al. Tumor necrosis factor-mediated disposition of infliximab in ulcerative colitis patients. *J Pharmacokinet Pharmacodyn*. 2019;46(6):543-551. doi:10.1007/s10928-019-09652-5

39.	Brandse JF, Mathôt RA, Kleij D van der, et al. Pharmacokinetic Features and Presence of Antidrug Antibodies Associate With Response to Infliximab Induction Therapy in Patients With Moderate to Severe Ulcerative Colitis. *Clinical Gastroenterology and Hepatology*. 2016;14(2):251-258.e2. doi:10.1016/j.cgh.2015.10.029

40.	Brandse JF, Mould D, Smeekes O, et al. A Real-life Population Pharmacokinetic Study Reveals Factors Associated with Clearance and Immunogenicity of Infliximab in Inflammatory Bowel Disease. *Inflamm Bowel Dis*. 2017;23(4):650-660. doi:10.1097/MIB.0000000000001043

41.	Dotan I, Ron Y, Yanai H, et al. Patient factors that increase infliximab clearance and shorten half-life in inflammatory bowel disease: a population pharmacokinetic study. *Inflamm Bowel Dis*. 2014;20(12):2247-2259. doi:10.1097/MIB.0000000000000212

42.	Dreesen E, Berends S, Laharie D, et al. Modelling of the relationship between infliximab exposure, faecal calprotectin and endoscopic remission in patients with Crohn’s disease. *Br J Clin Pharmacol*. 2021;87(1):106-118. doi:10.1111/bcp.14364

43.	Edlund H, Steenholdt C, Ainsworth MA, et al. Magnitude of Increased Infliximab Clearance Imposed by Anti-infliximab Antibodies in Crohn’s Disease Is Determined by Their Concentration. *AAPS J*. 2017;19(1):223-233. doi:10.1208/s12248-016-9989-8

44.	Fasanmade AA, Adedokun OJ, Blank M, Zhou H, Davis HM. Pharmacokinetic properties of infliximab in children and adults with Crohn’s disease: a retrospective analysis of data from 2 phase III clinical trials. *Clin Ther*. 2011;33(7):946-964. doi:10.1016/j.clinthera.2011.06.002

45.	Fasanmade AA, Adedokun OJ, Ford J, et al. Population pharmacokinetic analysis of infliximab in patients with ulcerative colitis. *Eur J Clin Pharmacol*. 2009;65(12):1211-1228. doi:10.1007/s00228-009-0718-4

46.	Grisic AM, Eser A, Huisinga W, Reinisch W, Kloft C. Quantitative relationship between infliximab exposure and inhibition of C-reactive protein synthesis to support inflammatory bowel disease management. *Br J Clin Pharmacol*. 2021;87(5):2374-2384. doi:10.1111/bcp.14648

47.	Kantasiripitak W, Verstockt B, Alsoud D, et al. The effect of aging on infliximab exposure and response in patients with inflammatory bowel diseases. *Br J Clin Pharmacol*. 2021;87(10):3776-3789. doi:10.1111/bcp.14785

48.	Magro F, Fernandes S, Patita M, et al. The Influence of Subclinical Active Inflammation on IFX Pharmacokinetic Modeling and Disease Progression Assessment: Findings from a Prospective Real-World Study in Inflammatory Bowel Disease Patients. *J Crohns Colitis*. 2024;18(7):1102-1112. doi:10.1093/ecco-jcc/jjae014

49.	Ternant D, Passot C, Aubourg A, et al. Model-Based Therapeutic Drug Monitoring of Infliximab Using a Single Serum Trough Concentration. *Clin Pharmacokinet*. 2018;57(9):1173-1184. doi:10.1007/s40262-017-0621-6

50.	Passot C, Mulleman D, Bejan-Angoulvant T, et al. The underlying inflammatory chronic disease influences infliximab pharmacokinetics. *MAbs*. 2016;8(7):1407-1416. doi:10.1080/19420862.2016.1216741

51.	Monolix 2024R1, Simulations Plus, doi: 10.5281/zenodo.11401936.

52.	Simulx 2024R1, Simulations Plus, doi: 10.5281/zenodo.11402004.

53.	Hansel J, Mannan F, Robey R, et al. Covariates in population pharmacokinetic studies of critically ill adults receiving β-lactam antimicrobials: a systematic review and narrative synthesis. *JAC Antimicrob Resist*. 2024;6(1):dlae030. doi:10.1093/jacamr/dlae030

54.	Roos JF, Bulitta J, Lipman J, Kirkpatrick CMJ. Pharmacokinetic-pharmacodynamic rationale for cefepime dosing regimens in intensive care units. *J Antimicrob Chemother*. 2006;58(5):987-993. doi:10.1093/jac/dkl349

55.	Jonckheere S, De Neve N, De Beenhouwer H, et al. A model-based analysis of the predictive performance of different renal function markers for cefepime clearance in the ICU. *J Antimicrob Chemother*. 2016;71(9):2538-2546. doi:10.1093/jac/dkw171

56.	Barreto EF, Chang J, Bjergum MW, et al. Adequacy of cefepime concentrations in the early phase of critical illness: A case for precision pharmacotherapy. *Pharmacotherapy*. 2023;43(11):1112-1120. doi:10.1002/phar.2766

57.	Seo H, Kim YK, Park S, Kim HI, Lee DH. Population Pharmacokinetics and Monte Carlo Simulation of Cefepime in Critically Ill Patients with Hospital-Acquired/Ventilator-Associated Pneumonia. *Infect Chemother*. 2023;55(1):29-41. doi:10.3947/ic.2022.0087

58.	Bilal M, Zoller M, Fuhr U, et al. Cefepime Population Pharmacokinetics, Antibacterial Target Attainment, and Estimated Probability of Neurotoxicity in Critically Ill Patients. *Antimicrob Agents Chemother*. 2023;67(7):e0030923. doi:10.1128/aac.00309-23

59.	Carlier M, Taccone FS, Beumier M, et al. Population pharmacokinetics and dosing simulations of cefepime in septic shock patients receiving continuous renal replacement therapy. *Int J Antimicrob Agents*. 2015;46(4):413-419. doi:10.1016/j.ijantimicag.2015.05.020

60.	Barreto EF, Chang J, Rule AD, et al. Population pharmacokinetic model of cefepime for critically ill adults: a comparative assessment of eGFR equations. *Antimicrob Agents Chemother*. 2023;67(11):e0081023. doi:10.1128/aac.00810-23

61.	An G, Creech CB, Wu N, et al. Population pharmacokinetics and target attainment analyses to identify a rational empirical dosing strategy for cefepime in critically ill patients. *J Antimicrob Chemother*. 2023;78(6):1460-1470. doi:10.1093/jac/dkad106

62.	Delattre IK, Musuamba FT, Jacqmin P, et al. Population pharmacokinetics of four β-lactams in critically ill septic patients comedicated with amikacin. *Clin Biochem*. 2012;45(10-11):780-786. doi:10.1016/j.clinbiochem.2012.03.030

63.	Green B, Duffull SB. What is the best size descriptor to use for pharmacokinetic studies in the obese? *Br J Clin Pharmacol*. 2004;58(2):119-133. doi:10.1111/j.1365-2125.2004.02157.x

64.	Wurtz R, Itokazu G, Rodvold K. Antimicrobial dosing in obese patients. *Clin Infect Dis*. 1997;25(1):112-118. doi:10.1086/514505

65.	Inker LA, Eneanya ND, Coresh J, et al. New Creatinine- and Cystatin C-Based Equations to Estimate GFR without Race. *N Engl J Med*. 2021;385(19):1737-1749. doi:10.1056/NEJMoa2102953

66.	Verdier MC, Tribut O, Tattevin P, Le Tulzo Y, Michelet C, Bentué-Ferrer D. Simultaneous Determination of 12 β-Lactam Antibiotics in Human Plasma by High-Performance Liquid Chromatography with UV Detection: Application to Therapeutic Drug Monitoring▿. *Antimicrob Agents Chemother*. 2011;55(10):4873-4879. doi:10.1128/AAC.00533-11
